# A PRISMA-Aligned Agentic Framework for Medical Systematic Reviews and Evidence Synthesis

**DOI:** 10.64898/2026.07.30.26359375

**Authors:** Haoming Huang, Qiaoyu Zheng, Pengcheng Qiu, Weike Zhao, Ya Zhang, Weidi Xie, Yanfeng Wang, Xiaoman Zhang, Chaoyi Wu

## Abstract

Medical systematic reviews are central to evidence-based medicine, but they remain slow, labor-intensive, and difficult to maintain under the full Preferred Reporting Items for Systematic Reviews and Meta-Analyses (PRISMA) workflow. Recent LLM-based deep research agents offer a promising route to addressing this challenge, yet reliable deployment in medical systematic reviews remains limited by insufficient clinical domain knowledge and inconsistent adherence to evidence-based methodological standards across the full workflow. We address these gaps with MedSR-Copilot, a PRISMA-aligned multi-agent copilot that decomposes review automation into literature retrieval, coarse-to-fine screening, data extraction, Risk-of-Bias assessment, and evidence synthesis, while preserving structured intermediate artifacts throughout the workflow. We further introduce MedSR-Bench, an end-to-end benchmark for evaluating systems beyond isolated subtasks, from review input to final evidence-synthesis conclusions. MedSR-Copilot completes medical systematic reviews end-to-end under the full PRISMA workflow, achieving 63.6% human-aligned conclusions, 18.3 percentage points above the best baseline among strong general-purpose LLMs and prior automated review systems. In a human-AI collaboration study involving 23 analysis groups across four systematic review topics, MedSR-Copilot, used as a copilot, reduces end-to-end review time by 64.9% and improves final conclusion accuracy by 27.4 percentage points compared with routine-practice workflows. Together, these results demonstrate the reliability and efficiency of MedSR-Copilot as a medical research copilot and suggest a practical path toward trustworthy review automation.

## 1 Introduction

*Systematic reviews* (SRs) and meta-analyses serve as the cornerstone of evidence-based medicine (EBM). By strictly identifying, evaluating, and synthesizing findings from cutting-edge clinical studies, they transform the latest research into gold-standard evidence that supports guideline updates, clinical practice, and the design of new trials or intervention studies [1, 2, 3, 4].

Despite their central role, producing high-quality SRs in a timely manner remains a major challenge. As shown in Fig. 1a, a rigorous PRISMA^1^-compliant review [5, 6] entails a series of complex, tightly coupled, and human-intensive steps, including literature retrieval, study screening, data extraction, and evidence synthesis, often taking over a year and requiring collaboration among multiple domain experts [7]. These manual steps are susceptible to fatigue, attention drift, and reviewer inconsistency, making it difficult to maintain an objective and reproducible evidence synthesis pipeline [8]. This already demanding process is further strained by the rapid growth of the biomedical literature: more than 1.5 million papers are added to PubMed annually, causing evidence updating to significantly lag and delaying their translation into patient care [9].

**Figure 1.**
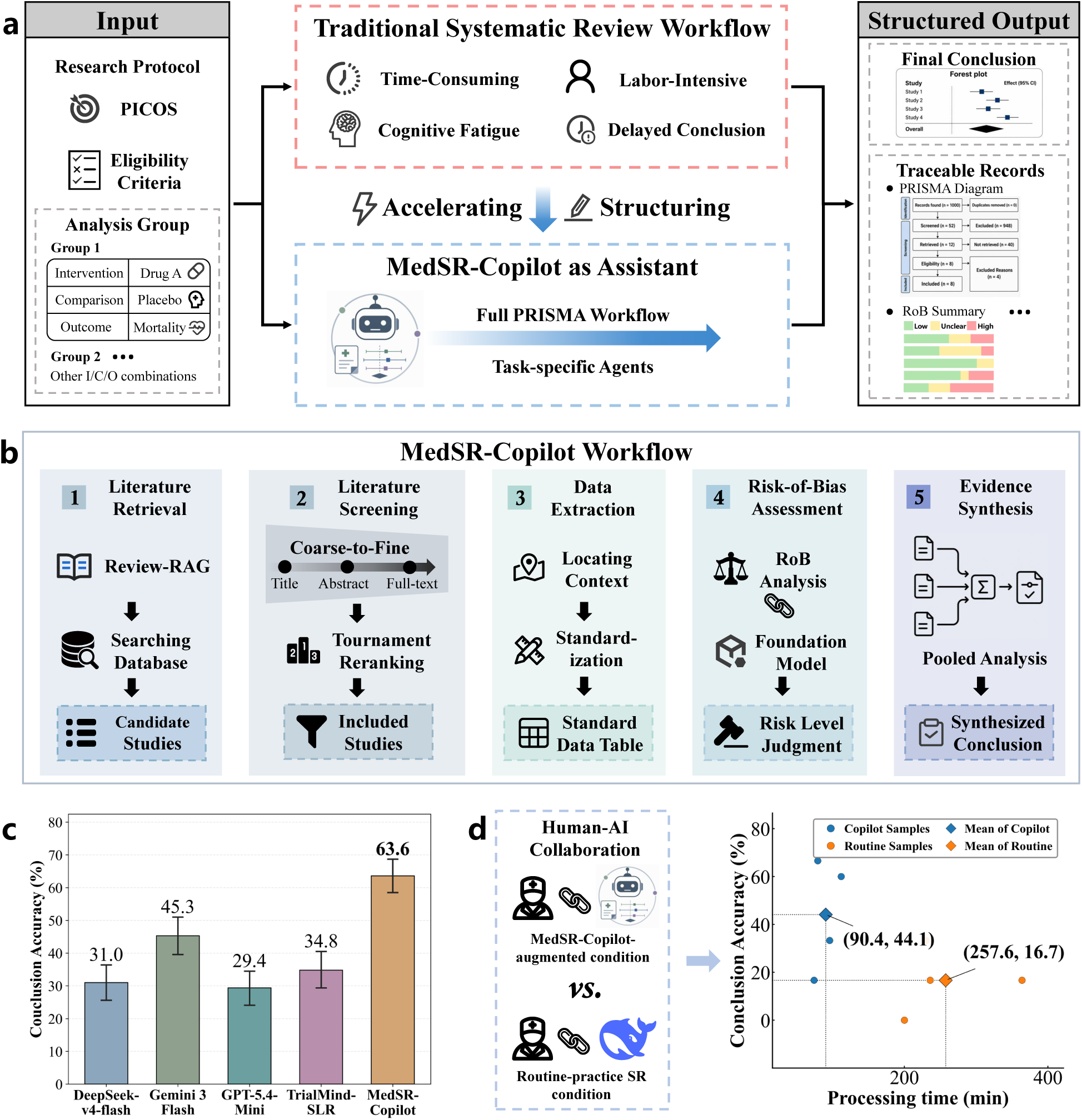
Overview of MedSR-Copilot and MedSR-Bench. **a**, Motivation for MedSR-Copilot. Medical systematic reviews take a research protocol as input and produce synthesized conclusions with traceable intermediate records. The traditional workflow is time-consuming, labor-intensive, slow to update, and prone to fatigue from repetitive tasks. MedSR-Copilot helps accelerate and structure this PRISMA-aligned process. **b**, System overview of MedSR-Copilot. The system is designed for literature retrieval, literature screening, data extraction, Risk-of-Bias assessment, and evidence synthesis through task-specific workflow components. **c**, End-to-end conclusion accuracy on a subset of MedSR-Bench (n=316), where MedSR-Copilot achieves the best final evidence-synthesis accuracy among the compared baselines. Error bars indicate 95% bootstrap confidence intervals. **d**, Human–AI collaboration results comparing conclusion accuracy and processing time. Compared with the routine-practice SR condition, the MedSR-Copilot-augmented condition achieves higher final conclusion accuracy with shorter processing time.

Recent LLM-based deep research agents [10, 11, 12] offer a promising route to addressing this challenge. By coupling iterative planning with search tools and cross-document synthesis, these systems have rapidly progressed from task-level assistance toward agentic support for end-to-end research workflows. They can tackle long-horizon scientific research tasks by automating literature exploration, aggregating evidence across sources, and synthesizing comprehensive literature reviews. However, their limitations in domain knowledge, workflow adaptation, and specialized tool use [13] constrain reliable deployment in domain-specific and methodologically rigorous workflows, with medical systematic reviews representing a particularly demanding case. These reviews require rigorous, evidence-grounded analysis within established clinical and methodological frameworks, ensuring that decisions concerning study inclusion, data extraction, and risk-of-bias assessment are well justified and traceable to the underlying evidence. Errors at any stage can propagate through subsequent steps and ultimately compromise the validity of the resulting synthesis.

To address this gap, we present **MedSR-Copilot**, a PRISMA-aligned agentic system for full-workflow support of medical systematic reviews and meta-analyses (Fig. 1b). Given a research protocol specifying PICOS^2^, eligibility criteria, and analysis-group definitions, MedSR-Copilot executes the entire PRISMA workflow and returns synthesized effect estimates and final conclusions with traceable intermediate action chains.

In its agentic architecture, MedSR-Copilot adopts task-specific designs to align each stage with expert review practice and improve workflow reliability. It comprises four task-specific agents for literature retrieval, literature screening, data extraction, and Risk-of-Bias (RoB) assessment, together with a dedicated engine for final evidence synthesis, thereby supporting the **full PRISMA workflow**. For retrieval and screening, MedSR-Copilot combines a Review-RAG module with a coarse-to-fine, three-level hierarchical filtering pipeline, mirroring how reviewers derive search keywords from prior studies and progressively narrow candidates from broad relevance checks to fine-grained eligibility assessment. For data extraction, MedSR-Copilot adopts a two-stage paradigm that first locates supporting source contexts and then converts them into standardized meta-analytic data, reducing the LLM’s context burden and improving extraction reliability. For RoB assessment, which requires reasoning over professional RoB criteria, we develop SR-RoB-7B, a dedicated model enhanced with reinforcement-learning-based reasoning training for domain-level bias judgment. Under this PRISMA-aligned design, each subtask increases task-specific reasoning effort, improves traceability, and reduces error propagation, thereby ensuring the generation of reliable synthesized conclusions.

To evaluate MedSR-Copilot, we first consider four existing public benchmarks: TrialReviewBench [14], TrialPanorama [15], Yun Bench [16] (a data extraction benchmark introduced by Yun et al), and RoBBR [17]. However, existing benchmarks focus on only a part of subtasks within the SR pipeline, making it difficult to assess an agent’s capability for end-to-end evidence processing and synthesis. To address this limitation, we construct **MedSR-Bench**, a benchmark comprising **100** SRs across **24** medical domains, curated from Cochrane Library [18], which is an authoritative evidence-based medicine database. Each SR is annotated across the full PRISMA workflow, enabling both subtask-level process evaluation and end-to-end evaluation of final conclusion. In total, these annotations span **1,297** RCTs, **1,204** RoB-labeled samples, and **2,144** analysis groups, yielding **7,570** samples for data extraction and **1,908** samples for end-to-end conclusion evaluation.

In experiments, we evaluate MedSR-Copilot against 3 general LLMs (DeepSeek-v4-flash, Gemini 3 Flash, and GPT-5.4-Mini), 2 automated medical SR systems, and a task-specific retrieval model across end-to-end and subtask-level settings as applicable. MedSR-Copilot demonstrates consistent improvements. In end-to-end evaluation on MedSR-Bench, MedSR-Copilot shows strong reliability in automated research, producing SR conclusions aligned with human experts in 63.6% of cases and outperforming both LLM baselines and automated SR systems, with an 18.3 percentage-point improvement over the second-best method (Fig. 1c). In subtask evaluation, all specialized sub-agents in MedSR-Copilot improve their corresponding metrics over prior baselines across PRISMA subtasks. In particular, for literature screening, it achieves the highest F1 score across all three benchmarks, indicating a stronger balance between eligible-study retention and false-positive exclusion. Furthermore, we conduct human-AI collaboration testing. Compared with routine-practice SR workflows, where human participants could use everyday tools such as general LLM assistance and web search, adding MedSR-Copilot to their workflow improves final conclusion accuracy from 16.7% to 44.1% and reduces total processing time by 64.9%. These results demonstrate its practical value as an expert-facing copilot (Fig. 1d).

## 2 Results

In this section, we present our main results, beginning with a brief overview of MedSR-Copilot and the experimental setup.

### 2.1 MedSR-Copilot overview

MedSR-Copilot is designed to automatically support the full medical SR workflow from a research protocol input to final synthesized conclusions. Throughout this process, users may optionally review and revise intermediate results as part of a human-in-the-loop workflow.

Specifically, to initiate an SR, users first input a structured research protocol including three parts: a PICOS-based review question, eligibility criteria, and predefined analysis groups. The eligibility criteria define which studies should be included or excluded in literature screening. Because a single research topic may contain multiple specific interventions, comparisons, and primary or secondary outcomes, an analysis group is defined as a specific intervention–comparison–outcome unit under the review topic and serves as the basic unit for data extraction and evidence synthesis. For example, under the comparison of combination therapy versus radiotherapy, progression-free survival as the primary outcome and tumor area as the secondary outcome define two separate analysis groups.

MedSR-Copilot then invokes a series of specialized subagents to complete the full workflow. The ‘literature retrieval’ subagent first retrieves candidate clinical studies based on PICOS, and the ‘literature screening’ subagent filters them according to the eligibility criteria, producing a user-refinable list of included clinical study papers. Then, given the user-confirmed paper list and predefined analysis groups, the ‘data extraction’ and ‘Risk-of-Bias assessment’ subagents extract evidence and assess bias risk of included studies, respectively. The extracted data can also be optionally inspected and revised by users. Finally, they are passed to the ‘evidence synthesis’ module, which finalizes the outputs, including synthesized effect estimates, statistical results, forest plots, and conclusions for each analysis group. To improve transparency and auditability, MedSR-Copilot also records intermediate results as traceable reasoning process output, such as the generated PRISMA flow diagram, structured extraction tables, and RoB summary plots.

### 2.2 Experiment Setup

In this part, we briefly introduce the experimental setup, including benchmark datasets, comparison baselines, and evaluation metrics.

**Benchmark datasets.** We adopt five benchmarks for evaluation, consisting of four public datasets and our proposed MedSR-Bench. The public datasets are primarily used to assess the performance of individual subagents. In particular, TrialReviewBench [14] and TrialPanorama [15] are used for evaluating paper retrieval and screening, Yun Bench [16] is used for data extraction, and RoBBR [17] is used for RoB assessment. Our proposed MedSR-Bench is used for both subagent-level and end-to-end evaluation, where the latter assesses whether the final SR conclusions end-to-end generated by MedSR-Copilot through the multi-step pipeline are consistent with human-produced results. Notably, during evaluation, all methods are restricted to using clinical studies published before the corresponding SR test sample, mirroring the information available to human reviewers at the time. Thus, the SR’s content can be viewed as a human expert reference ground truth.

**Baseline methods.** We compare MedSR-Copilot with three categories of baselines:

- **General LLMs.** DeepSeek-v4-flash, Gemini 3 Flash, and GPT-5.4-Mini are evaluated on all subtasks using task-specific prompts tailored to each specialized setting. For the end-to-end evaluation, we further chain these task-specialized prompts together to form a straightforward baseline agent harness, enabling automated SR execution.
- **Task-specialized models.** DeepRetrieval [19], a specialized model for search-query rewriting, is used as the retrieval-specific baseline.
- **Automated systematic-review agents.** We consider two existing SR agents, Manalyzer [20] and TrialMindSLR [14], for comparison. Manalyzer [20] is a general meta-analysis system and is only considered for retrieval and screening evaluation as it does not cover other medical SR subtasks, like cohort data extraction and RoB assessment. TrialMindSLR [14] is an automated medical SR system, but its source code does not support final statistical synthesis. We therefore use the same ‘Statistical Synthesis’ Engine as MedSR-Copilot, enabling both end-to-end and subtask comparisons with it to assess the effectiveness of the other agentic modules.

**Evaluation metrics.** Different evaluation metrics are used for subtask and end-to-end evaluations. Specifically, literature retrieval is assessed using Recall@3000 and Recall@5000. Literature screening is primarily evaluated using F1 score, with Recall and Precision reported as complementary metrics based on comparisons between the final included-study list and the manually curated one. Data extraction is evaluated using Recall and Accuracy: Recall measures whether an analysis group is successfully identified, while Accuracy assesses whether the extracted numerical results are correctly standardized. Lastly, both RoB assessment and end-to-end evaluation are evaluated using Accuracy. For RoB assessment, Accuracy measures whether the predicted risk level (high, unclear, or low) matches the human-annotated label. For end-to-end evidence synthesis, Accuracy measures whether the predicted conclusion matches the human-derived conclusion. For example, a conclusive prediction is marked as correct if both the agent and human experts agree on whether a given intervention significantly reduces mortality compared with the control, under the same research protocol. The details of test set construction, baseline adaptation, and evaluation metrics can be found in **Method** section 4.3.

### 2.3 MedSR-Copilot Achieves the Highest Expert Alignment in Automated SR Research

In the end-to-end evaluation setting, MedSR-Copilot takes a research protocol as input, bypasses intermediate human-in-the-loop revision, automatically produces the final three-class evidence-synthesis conclusion for each predefined analysis group: favoring the intervention, favoring the control, or showing no significant effect. All baseline methods are evaluated under the same setting, and their final conclusion-level accuracy is reported against human-expert SR conclusions.

This end-to-end metric reflects their overall ability to complete the full SR pipeline, jointly capturing their effectiveness in evidence searching, extraction, and synthesis throughout the PRISMA-aligned SR workflow.

The results are shown in Figure 1c. MedSR-Copilot achieves the highest overall end-to-end conclusion accuracy of 63.6%, substantially outperforming TrialMindSLR (34.8%), DeepSeek-v4-flash (31.0%), Gemini 3 Flash (45.3%), and GPT-5.4-Mini (29.4%). End-to-end conclusion accuracy captures the accumulated deviation between an automated SR system and human reviewers after multiple pipeline stages. The higher accuracy of MedSR-Copilot suggests that it better controls error propagation across the SR workflow, enabling final conclusions that remain closely aligned with manual SRs.

We further regroup the test reviews into four broader medical domains and analyze their conclusion accuracy, respectively. As shown in Figure 2a, MedSR-Copilot consistently achieves the best performance in all four domains, achieving 62.1% in Maternal-Child, 61.3% in Internal, 61.1% in Surgical, and 81.2% in Alternative Medicine. Compared with the second-best baseline, MedSR-Copilot improves conclusion accuracy by 6.1, 23.3, 27.8, and 3.1 percentage points, respectively. These consistent gains suggest that MedSR-Copilot generalizes well across heterogeneous clinical areas rather than overfitting to a single domain.

**Figure 2.**
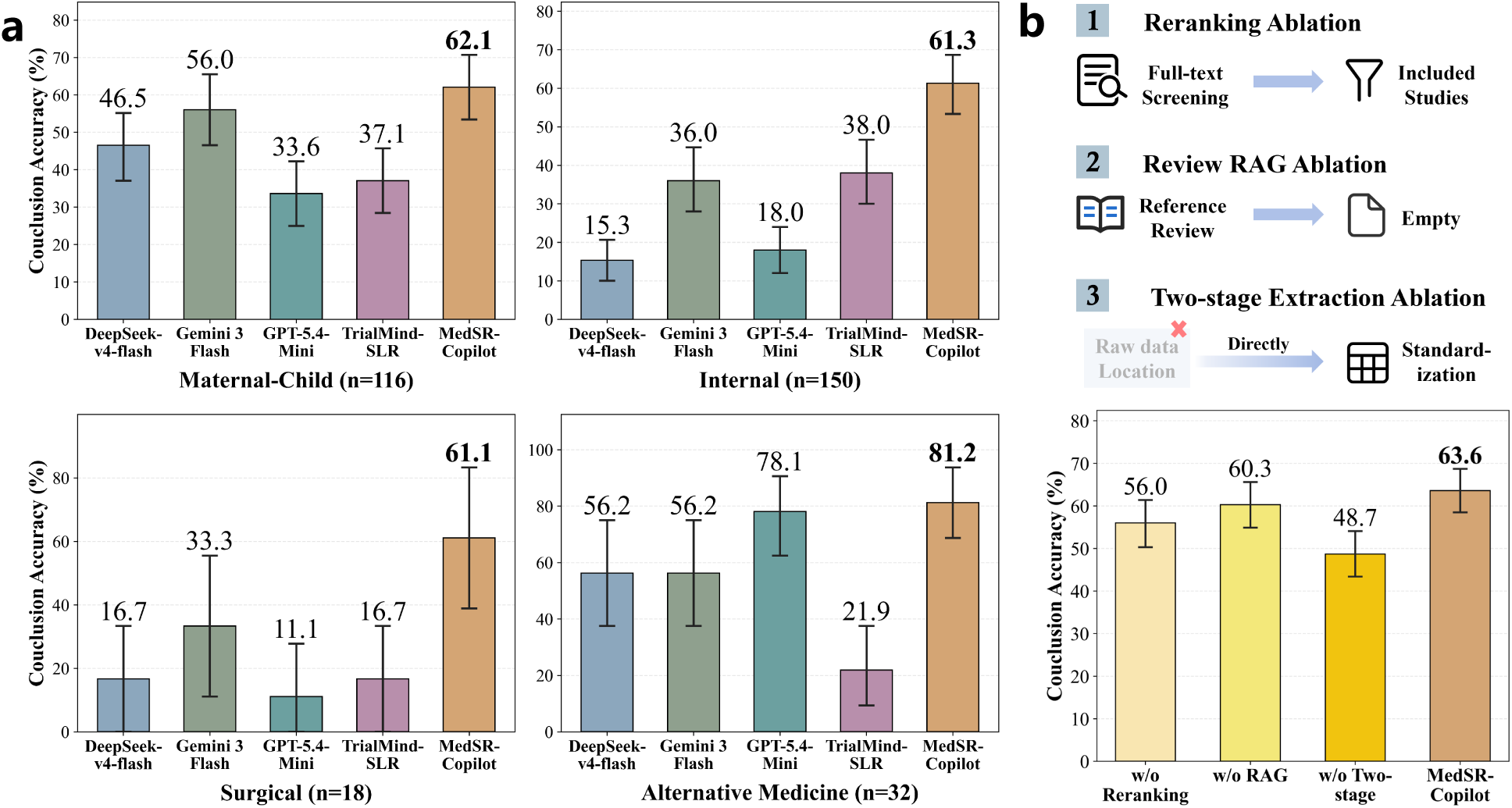
End-to-end conclusion evaluation on MedSR-Bench. **a,** Domain-level conclusion accuracy across four broad medical domains regrouped from the test subset. **b,** Ablation study of key MedSR-Copilot modules, including tournament reranking, Review-RAG, and two-stage data extraction. Conclusion accuracy is reported as percentage. Error bars indicate 95% bootstrap confidence intervals.

### 2.4 MedSR-Copilot Improves SR Accuracy and Efficiency in the Co-Researcher Setting

We conduct a human-AI collaboration experiment to evaluate whether MedSR-Copilot can support manual SR processes, where a human researcher paired with MedSR-Copilot as a co-researcher could outperform human researchers working with common tools.

We select four SR research protocols from MedSR-Bench, covering infectious disease, cardiovascular disease, occupational health, and health systems, respectively. Each review contains five to six analysis groups, yielding 23 analysis-group-level conclusion samples and 110 data-extraction samples in total. For each protocol, we construct a closed candidate-study pool to prevent participants from searching for newly published studies or accessing the original published SR, ensuring reproducibility. Specifically, each pool included a sampled subset of ground-truth studies from the source review, supplemented with non-ground-truth studies retrieved during the corresponding literature search, resulting in 200 candidate studies per topic. Experts are instructed to perform strict medical SR within the provided candidate-study pool.

The experiment compares two conditions: a routine-practice condition and a MedSR-Copilot-augmented condition. In the routine-practice condition, participants follow their usual research workflow and are allowed to use any familiar tools, like general-purpose LLMs, web search, reference management tools, and other standard resources. In the MedSR-Copilot-augmented condition, experts follow the same routine-practice workflow with additional access to the MedSR-Copilot platform.

Four participants with medical backgrounds are assigned using a balanced crossed design. All participants provided informed consent before taking part in the study. Each participants completes two distinct SRs, one under the routine-practice condition and the other under the MedSR-Copilot-augmented condition, such that each of the four selected topics is completed twice, once under each condition, by different participants. This assignment avoided repeated exposure of the same participant to the same topic while enabling within-topic comparison between the two conditions. For comparison, we evaluate screening recall and precision, data-extraction recall and accuracy, end-to-end conclusion accuracy, and the time required to complete each review.

As shown in Figure 1d, the MedSR-Copilot-augmented condition achieved higher end-to-end conclusion accuracy with less processing time than the routine-practice SR condition. Final conclusion accuracy increased from 16.7% in the routine-practice SR condition to 44.1% in the MedSR-Copilot-augmented condition, while total processing time decreased by 167.2 minutes, corresponding to a 64.9% reduction. These findings suggest that MedSR-Copilot provides a structured intermediate SR reasoning process and PRISMA-aligned task decomposition, helping researchers make more accurate synthesis decisions with less manual effort. Clinically, such gains could reduce a key bottleneck in evidence generation: the time and expertise required to produce reliable systematic reviews that inform guidelines, health technology assessments, and policy decisions. By improving conclusion-level accuracy while reducing review time, MedSR-Copilot may support faster and more reproducible evidence synthesis, especially for rapidly evolving clinical questions. More detailed results for the screening and data extraction subtasks, together with the system usability evaluation, are presented in Appendix A.1.1 and Appendix A.1.2.

### 2.5 MedSR-Copilot Outperforms Related Baselines Across All SR Subtasks

In addition to the whole-system-level evaluations, we perform subtask-level comparisons to assess the effectiveness of our subagent designs. Notably, because the final evidence synthesis step is a fixed heuristic procedure applied to previously processed outputs and does not require a specialized agentic design, we do not report it as a separate subtask.

**Literature retrieval results.** For literature retrieval, the agent takes the ‘Population’ and ‘Intervention’ components of PICOS as input and returns a ranked list of candidate studies. MedSR-Copilot uses the Review-RAG module to retrieve related systematic reviews as references and then refines the initial search queries through query generation and augmentation. During evaluation, Review-RAG only accesses reviews published before the corresponding source SR.

Across the three benchmarks, MedSR-Copilot achieves the best retrieval recall on most metrics (Figure 3). Specifically, on TrialReviewBench, MedSR-Copilot obtains Recall@3000 of 74.8% and Recall@5000 of 79.6%; on TrialPanorama, it obtains Recall@3000 of 81.4% and Recall@5000 of 84.2%; and on MedSR-Bench, it obtains Recall@3000 of 78.1% and Recall@5000 of 82.2%. Compared with TrialMindSLR, another automated system specialized for medical systematic reviews, MedSR-Copilot improves Recall@3000 by 2.5, 10.2, and 7.9 percentage points across the three datasets, respectively, and improves Recall@5000 by 3.1, 11.7, and 8.4 percentage points. The ablation analysis of the Review-RAG module is provided in Appendix A.1.4. These gains indicate that Review-RAG provides useful review-level context for query construction, improving the recall of eligible studies.

**Figure 3.**
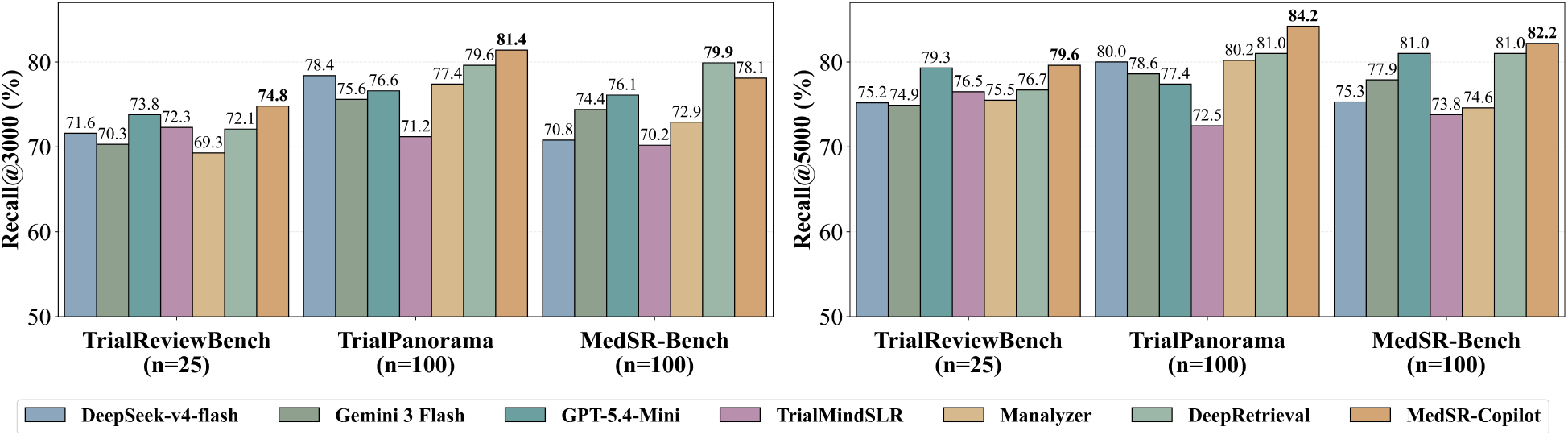
Retrieval performance on TrialReviewBench, TrialPanorama, and MedSR-Bench. Recall@3000 and Recall@5000 are reported as percentages.

**Literature screening results.** For literature screening, given a candidate paper set, MedSR-Copilot filters them based on the input PICOS and eligibility criteria, then outputs the final included-study list. MedSR-Copilot follows a fully LLM-based, coarse-to-fine PRISMA-aligned screening process, covering title-, abstract-, and full-text-level screening. At the title and abstract levels, candidate studies are assessed according to PICOS requirements. At the full-text level, detailed inclusion and exclusion criteria are further used for eligibility assessment. After full-text screening, a tournament-inspired reranking module [21] ranks the passed studies, and MedSR-Copilot retains the top-*k* studies as final candidates. We set *k* = 20 in all screening experiments. Such a progressive agentic screening design ensures that each step operates within the LLM’s effective context window, fully leveraging the superior semantic understanding capabilities of the latest LLMs.

The results in Table 1 show that MedSR-Copilot achieves the highest F1 score on all three datasets, reaching 46.3%, 43.9%, and 51.0%, respectively. It also achieves the highest precision, while maintaining competitive recall among the other two automated systematic-review systems. In contrast, Gemini 3 Flash achieves the highest recall but substantially lower F1 scores of 22.7%, 19.3%, and 19.3% across the three datasets, indicating that it tends to include studies from the input list indiscriminately. These results demonstrate that MedSR-Copilot provides a stronger overall balance between retaining eligible studies and excluding false positives, thereby reducing noisy evidence passed to downstream stages. An ablation analysis of the full-text reranking module is provided in Appendix A.1.5.

**Table 1.**
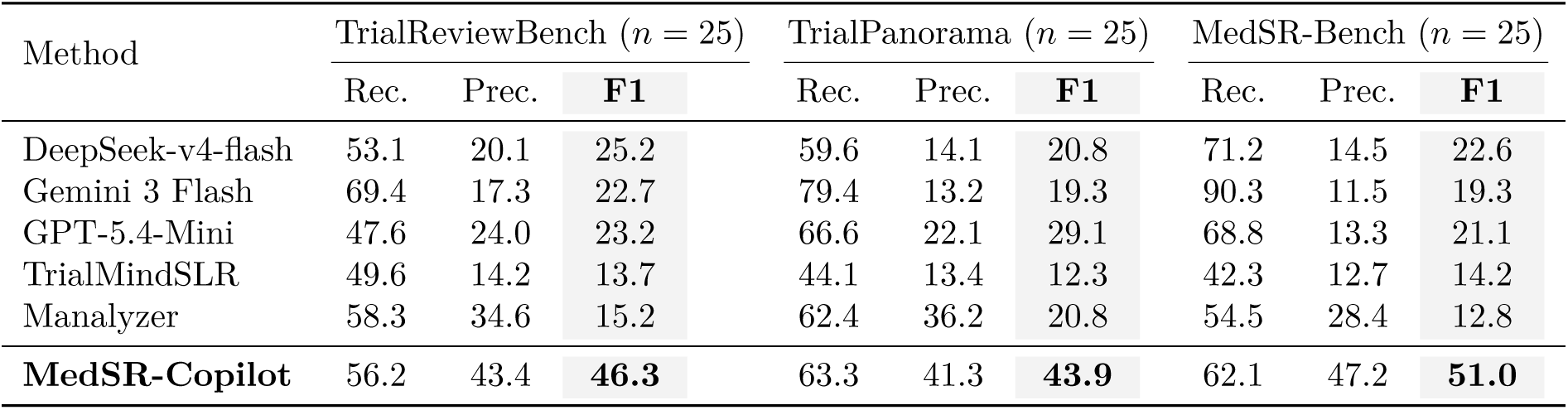
Literature screening performance on TrialReviewBench, TrialPanorama, and MedSR-Bench. F1 score (F1) is the primary metric, with Recall (Rec.) and Precision (Prec.) reported as complementary metrics. All metrics are reported in percentage points.

**Data extraction results.** For data extraction, the input consists of the full texts of ground-truth included studies and the predefined analysis-group information. MedSR-Copilot decomposes the task into a two-stage agentic workflow: it first focuses on locating the source text and raw data of each analysis group, and then converts the extracted source text into standardized numerical data as task outputs, thereby reducing the burden of handling evidence localization and data standardization simultaneously.

As shown in Figure 4a, MedSR-Copilot achieves the strongest data-item accuracy on both MedSR-Bench and Yun Bench, while maintaining high group recall. On MedSR-Bench, MedSR-Copilot achieves the best performance on both metrics, with group recall of 90.5% and data-item accuracy of 63.5%. On Yun Bench, it obtains competitive group recall of 97.6% and the highest data-item accuracy of 63.4%. These results show that the two-stage extraction workflow improves the accuracy of numerical data extraction and standardization while preserving strong analysis-group localization capability.

**Figure 4.**
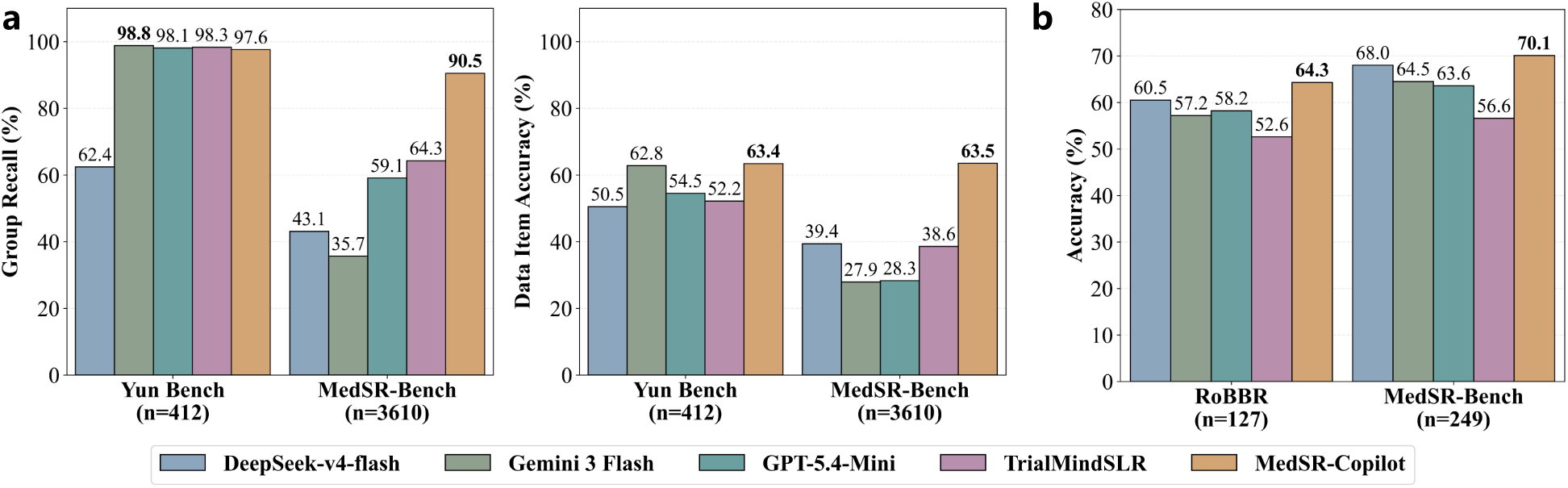
Data extraction and RoB assessment performance. **a,** Data extraction performance on Yun Bench and MedSR-Bench. Group recall and data-item accuracy are reported as percentages. **b,** Average Risk-of-Bias assessment performance on RoBBR and MedSR-Bench. Judgment accuracy is reported as the average across all samples.

**Risk-of-Bias assessment results.** For RoB assessment, the task takes the full text of each included study as input, and outputs risk judgments for five bias domains in the original Cochrane Risk-of-Bias tool for randomized trials, which we refer to as RoB 1 tool [22]. MedSR-Copilot develops a new SR-RoB-7B reasoning language model to produce domain-level judgments with supporting evidence, enabling better understanding of the RoB criteria.

As shown in Figure 4b, MedSR-Copilot with SR-RoB-7B achieves the best average judgment accuracy on both RoBBR and MedSR-Bench, reaching 64.3% and 70.1%, respectively. Compared with the second-best baseline, DeepSeek-v4-flash, SR-RoB-7B improves accuracy by 3.8 and 2.1 percentage points on the two datasets, respectively. These results indicate that task-specific training improves the reliability of RoB assessment. Detailed analyses across the five RoB domains are provided in Appendix A.1.6.

### 2.6 Ablation Study Confirms the Contribution of Each Agentic Design in MedSR-Copilot

We conduct ablation experiments in the end-to-end setting to examine the contributions of three key agentic designs in MedSR-Copilot: Review-RAG for literature retrieval, tournament-style reranking for literature screening, and the two-stage workflow for data extraction. Notably, the ‘RoB assessment’ module provides a complementary analysis of bias risk for the final conclusion, but does not affect the predicted conclusion category. Therefore, we do not ablate it. Implementation details of these ablation variants are provided in Appendix A.1.3.

The ablation results show that all three modules contribute to final conclusion accuracy (Figure 2b). Removing tournament reranking, Review-RAG, and two-stage data extraction reduces overall accuracy by 7.6, 3.3, and 14.9 percentage points, respectively. The largest decline comes from removing two-stage extraction, suggesting that explicitly locating source evidence before standardizing numerical data is critical for preventing extraction errors from propagating into final evidence synthesis.

Ablation studies of Review-RAG in the literature retrieval subtask evaluation and the reranking module in literature screening are provided in Appendix A.1.4 and Appendix A.1.5, respectively.

## 3 Discussion

In this work, we propose MedSR-Copilot, an agentic copilot designed to support medical systematic reviews with full PRISMA workflow coverage. To support more comprehensive evaluation in this area, we further introduce MedSR-Bench. By aligning with standard manual SR procedures, preserving transparent and auditable intermediate records, and enabling flexible human-in-the-loop revision, MedSR-Copilot reduces the manual workload of evidence synthesis while improving review quality.

### A medical SR agentic copilot for full PRISMA workflow coverage

General deep research agents are increasingly capable of planning, retrieval, and tool use for broad research tasks, but they are not inherently designed for fixed, protocol-governed medical SR workflows. SR-oriented agentic systems move closer to this setting by supporting literature search, screening, evidence extraction, or synthesis [14, 23, 24, 25]. However, they usually automate selected SR stages without fully reproducing the standard multi-stage human review practice across the full PRISMA pipeline. Moreover, automating a medical SR is not simply a matter of chaining workflow steps, because the reliability of final evidence synthesis can be constrained by the weakest stage. Each step requires careful reasoning and reflection to preserve methodological quality and maintain auditable PRISMA records. This partial workflow alignment and limited subtask-specialized reasoning can weaken evidence traceability and amplify cross-stage errors when generating final conclusions. Against this background, MedSR-Copilot contributes a medical SR copilot designed around the full PRISMA workflow rather than isolated automation modules, integrating the complete review process from study identification to evidence synthesis with traceable intermediate records.

### Task-specialized agentic designs improve subagent performance

To improve SR subtask performance, we propose diverse agentic designs. In retrieval, MedSR-Copilot uses Review-RAG to leverage related reviews as references for query construction, better matching how human reviewers scope a topic before formal search. In screening, MedSR-Copilot adopts a coarse-to-fine process with title-, abstract-, and full-text-level decisions, followed by full-text reranking to better distinguish highly similar studies under detailed eligibility criteria. For later stages, MedSR-Copilot also follows task-specific designs, including two-stage extraction and a dedicated RoB reasoning language model, to better support structured evidence processing. Different from general literature reviews, medical SRs require more precise, rigorous, and fine-grained reasoning. Our specialized agentic design explicitly targets these demands by decomposing the SR workflow into clinically grounded subtasks and enforcing stage-specific agentic workflows, thereby strengthening fine-grained evidence extraction and reasoning across the SR process.

### A new SR benchmark for end-to-end evaluation of automated systematic review systems

Our work also introduces MedSR-Bench, a benchmark designed for evaluation beyond isolated subtasks. Unlike prior resources that mainly focus on only part of the subtasks, MedSR-Bench supports both subtask-level evaluation and end-to-end evaluation of final synthesized conclusions under the full PRISMA workflow. The benchmark contains 100 systematic reviews spanning 24 medical domains, providing a broader basis for studying cross-stage error propagation and conclusion-level reliability in automated SR systems.

### MedSR-Copilot as an effective AI co-researcher for SR

Beyond automated evaluation, our results suggest that MedSR-Copilot can serve as an effective AI co-researcher integrated into practical SR workflows. In the human-AI collaboration setting, the MedSR-Copilot-augmented condition improves final conclusion accuracy from 16.7% to 44.1% while reducing total review time by 167.2 minutes. These results indicate that PRISMA-aligned decomposition and structured intermediate outputs can improve both the quality and efficiency of researcher-led evidence synthesis. More broadly, they suggest that a workflow with clear intermediate records can help experts inspect, verify, and revise key decisions during the review process, rather than relying only on a final answer.

### Limitations and Future Development

Despite these advancements, MedSR-Copilot still has limitations. Its retrieval module is currently limited to PubMed, and future versions should incorporate broader databases such as Embase [26] and CENTRAL [27]. The system is also mainly designed for interventional systematic reviews, and should be extended to diagnostic and prognostic reviews. In addition, RoB judgment accuracy and data-item extraction accuracy still need further improvement to support more reliable evidence synthesis.

## 4 Methods

In this section, we first present the problem formulation that defines the full PRISMA workflow. We then describe MedSR-Bench, a benchmark curated from Cochrane reviews that serves as evaluation infrastructure and provides RoB-labeled training data for SR-RoB-7B, and the design of MedSR-Copilot, a PRISMA-aligned [6] agentic system that uses Gemini 3.1 Flash-Lite for most workflow components and SR-RoB-7B as the task-specific model for RoB assessment.

### 4.1 MedSR-Copilot Designs

In this section, we detail the design of MedSR-Copilot, an LLM-based agentic system for automated medical systematic reviews.

Overall, MedSR-Copilot coordinates specialized subagents and tools through a PRISMA-aligned agentic workflow. First, the retrieval subagent returns a list of candidate PubMed records, which is progressively refined by the screening subagent. Based on the resulting list of included studies, the extraction subagent then produces standardized numerical evidence for evidence synthesis, while the RoB subagent generates the PRISMA-required methodological judgments for the included studies. Throughout the workflow, MedSR-Copilot maintains structured intermediate artifacts at each stage, thereby supporting human review and revision of any intermediate results. The software-level platform implementation of MedSR-Copilot is described in Appendix A.3.

In our implementation, the RoB assessment module uses the task-specific SR-RoB-7B model, whereas all other workflow components of MedSR-Copilot are implemented with Gemini 3.1 Flash-Lite.

#### 4.1.1 Problem Formulation

We formulate medical systematic review as a research problem in which a structured review protocol is transformed into analysis-group-level research conclusions based on evidence from the existing literature.

We denote the input SR research protocol as

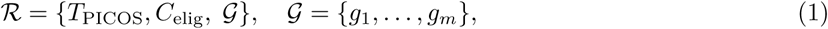

where *T*_PICOS_ is the PICOS-based research question, *C*_elig_ denotes the eligibility criteria, and G denotes the predefined analysis groups.

Our proposed MedSR-Copilot consists of four subagents, i.e., literature retrieval, literature screening, data extraction, and RoB assessment, and can be denoted as:

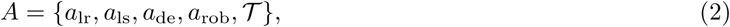

where *a*_lr_, *a*_ls_, *a*_de_, and *a*_rob_ denote the literature retrieval, literature screening, data extraction, and RoB assessment agents, respectively, powered by LLMs, and T denotes the external tool set available for the agents to call. The final evidence synthesis is implemented by a dedicated synthesis engine in T , rather than by an LLM-powered agent.

Given the input protocol *R*, MedSR-Copilot first produces traceable intermediate results:

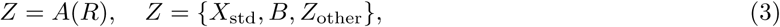

where *X*_std_ denotes the standardized study-level numerical evidence extracted for downstream statistical synthesis, B denotes the RoB judgments and supporting evidence, and *Z*_other_ denotes the other intermediate records, such as study-selection flow records, PRISMA flow diagrams, and RoB summary plots.

The final analysis-group-level synthesized outputs are then produced by the statistical synthesis engine:

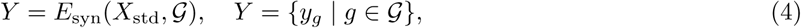

where *E*_syn_ denotes the final evidence-synthesis engine, which is implemented as a fixed statistical algorithm rather than an LLM-powered agent. Each *y_g_* ∈ *Y* includes the synthesized effect, statistical results, forest plot, and synthesized conclusion for analysis group *g*. Therefore, the overall system output is (*Y, Z*), where *Y* represents the final synthesized conclusions and related statistical outputs, while *Z* represents the PRISMA-required intermediate results. RoB assessment results are retained as part of *Z*, but only *X*_std_ is used as the input to *E*_syn_ for generating *Y* .

We then introduce the implementation of MedSR-Copilot, A = {*a*_lr_*, a*_ls_*, a*_de_*, a*_rob_, T }, formulating and clarifying the designs of its external tool set and four LLM-based agents.

#### 4.1.2 System Tools

Before introducing the LLM-based agents, we first define the tool set T that agents can invoke within their workflows.

**NCBI Entrez Literature Access Module.** We use the NCBI Entrez Programming Utilities (E-utilities)

[28] as the programmatic interface to PubMed. In the literature retrieval stage, the module takes the generated PubMed search query as input and obtains the corresponding PMID (PubMed Identifier) list returned by the PubMed search engine. In the screening stage, it takes PMID lists as input and retrieves the titles and abstracts of the corresponding articles.

**Full-text Content Pool.** We construct a dynamically updated full-text content pool of clinical studies. This pool contains the full texts of ground-truth studies in MedSR-Bench as well as non-ground-truth studies that are used for full-text screening. We parse these full texts into textual content with MinerU. During full-text screening, data extraction, and RoB assessment, MedSR-Copilot directly retrieves available study texts from this pool.

**Visualization Engine.** We implement a visualization engine to generate the visual outputs required by the SR workflow. Based on the number of records retained at each retrieval and screening stage, the engine draws the PRISMA flow diagram. Based on the distribution of RoB judgments across included studies, it draws RoB bar plots. Based on study-level data and pooled effects for each analysis group, it draws forest plots.

**Statistical Synthesis Engine.** We implement a statistical synthesis engine for evidence synthesis. The engine applies a random-effects model to the standardized study-level data extracted for each analysis group. Given the numerical results of individual studies, it outputs the pooled effect estimate, confidence interval, and heterogeneity statistics, which are then used to derive the final synthesized conclusion.

#### 4.1.3 Literature Retrieval Agent

We formulate literature retrieval as a candidate-study identification task from the structured review question. The literature retrieval module is defined as

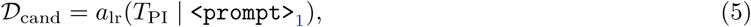

where *a*_lr_ denotes the literature retrieval agent,

<PROMPT>_1_ denotes its prompt, and D_cand_ denotes the retrieved candidate-study set. Following the practice of human experts conducting systematic reviews, *a*_lr_ only uses *T*_PI_, the Population and Intervention components of PICOS terms as the primary retrieval input [5], since these components are sufficient to determine the review topic while minimizing search restrictions and preserving broad recall.

To provide domain-specific knowledge and reference search strategies for query construction, MedSR-Copilot incorporates a review-grounded retrieval-augmented generation component, termed the **Review-RAG module**, within the literature retrieval agent. Specifically, given the Population and Intervention components as search inputs, Review-RAG retrieves related systematic reviews from PubMed with accessible PMC full texts and uses their full-text contents as references for query generation.

Based on *T*_PI_ and the retrieved review references, MedSR-Copilot generates PubMed search terms through a structured two-step process: initial generation of relevant medical subject headings (MeSH) terms [29] and free-text keywords, followed by augmentation through synonym replacement and decomposition of compound elements. The final query components are constructed by taking the union of the initial and augmented terms.

The generated terms are then converted into a Boolean PubMed query. Multiple terms within each category are combined using the “OR” operator to form distinct term groups, and the Population and Intervention groups are joined using the “AND” operator. Using the NCBI Entrez Literature Access Module, MedSR-Copilot submits the query to PubMed and obtains the returned PMID list, which forms D_cand_ and is passed to the hierarchical screening module.

#### 4.1.4 Hierarchical Literature Screening Agent

We formulate literature screening as a study selection task that maps the retrieved candidate-study set to the final included-study set under the predefined eligibility criteria. Generally, the literature screening agent can be formulated as:

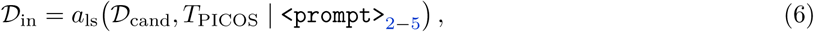

where *a*_ls_ denotes the literature screening agent, D_in_ denotes the final studies retained after the filtering,

<PROMPT>_2*−*5_ denotes the related prompts used in the process.

Specifically, following the PRISMA framework, MedSR-Copilot implements *a*_ls_ as a hierarchical coarse-to-fine screening pipeline, including title-level, abstract-level, and full-text-level screening followed by reranking. The NCBI Entrez Literature Access Module is used to obtain titles and abstracts for studies in D_cand_, while available full texts are retrieved from the full-text content pool. The title and abstract stages use PICOS terms *T*_PICOS_ as screening criteria, whereas the full-text stage further incorporates detailed inclusion and exclusion criteria from *C*_elig_.

Screening at all three levels is formulated as a classification problem. For each criterion, the model provides one of three judgments: “consistent”, indicating that the text explicitly contains matching or synonymous information; “inconsistent”, indicating the explicit presence of contradictory information; and “uncertain”, which is used when no consistent or sufficiently relevant information appears.

**Title-level Screening.** This stage evaluates the titles of all retrieved studies. It uses only the Population (P) and Intervention (I) terms from PICOS as screening criteria and applies the broadest inclusion standard. A study passes this stage if the judgments for both P and I are not “inconsistent”, formulated as:

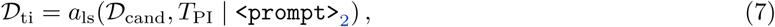

where D_ti_ denotes the studies retained after title-level screening, *T*_PI_ denotes the Population and Intervention components of *T*_PICOS_.

**Abstract-level Screening.** The input at this stage includes the abstract texts of the studies passing title-level screening and all five PICOS terms as screening criteria. A study is preliminarily included if the assessments for the P, I, and S dimensions are not all “uncertain” and contain no “inconsistent” ratings.

To maintain a manageable volume for the computationally expensive full-text review, MedSR-Copilot ranks these preliminarily included studies. MedSR-Copilot calculates a cumulative score for each study across the five PICOS dimensions by assigning “+1” to “consistent”, “-1” to “inconsistent”, and “0” to “uncertain”, with the weights for the core P and I dimensions doubled. The top 100 studies, including ties, pass to the full-text screening stage, formulated as:

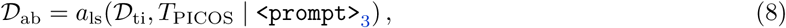

where D_ab_ denotes the studies retained after abstract-level screening.

**Full-text-level Screening.** The final screening stage evaluates the full texts of the studies that passed abstract screening. Because full texts provide more comprehensive details, we supplement the base PICOS with detailed inclusion and exclusion criteria for the P, I, and O terms, such as specific age limits or intervention sub-categories. At this stage, MedSR-Copilot applies rigorous standards: all five components of the criteria must be rated as “consistent” for preliminary inclusion, formulated as:

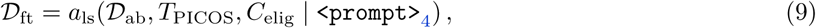

where D_ft_ denotes the studies retained after full-text-level screening.

**Tournament-inspired Full-text Reranking.** To further select high-quality studies from the preliminarily included full-text candidates, MedSR-Copilot employs a tournament-inspired reranking strategy. Tournament ranking mitigates the inefficiency of pairwise full-text comparisons and the context-length constraints of listwise ranking by partitioning candidates into small groups that can be evaluated in parallel [30, 31], formulated as:

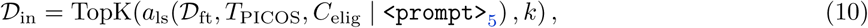

where D_in_ denotes the final included-study set obtained by retaining the top-*k* studies after full-text reranking. Our approach initiates *R* = 8 concurrent tournament instances. Within each stage of a tournament, candidate studies are partitioned into groups of 5. MedSR-Copilot evaluates the 5 full texts within a group and selects the top 2 studies to advance to the next round, with each advancement contributing one point to the study’s score. This elimination process continues until the final top 2 studies are determined for each tournament. To improve robustness, we shuffle the initial groupings across the 8 parallel tournaments. A study’s final reranking score is the cumulative number of advancement points across all tournaments. All studies are ranked by this score, and the top *k* studies, where *k* is user-specified with a default value of 20, are retained as the final included studies. The details about tournament reranking can be found in Appendix A.2.1.

At the end of the screening module, the visualization engine draws the PRISMA flow diagram based on the number of records retained at each stage.

#### 4.1.5 Data Extraction Agent

We formulate data extraction as a structured evidence extraction task from included studies to standardized study-level numerical results for predefined analysis groups. For each included study in D_in_, MedSR-Copilot retrieves the full text from the full-text content pool and extracts the outcome data associated with the target analysis groups. Specifically, *a*_de_ follows a two-stage design: localization and source-context extraction, followed by numerical standardization.

**Analysis-group Localization and Source-context Extraction.** In the first stage, *a*_de_ performs analysis-group localization and source-context extraction. Given the final included-study set and the predefined analysis groups, it locates the corresponding evidence in the full text and outputs the source context and raw data that support each standardized data item. This step explicitly links each required numerical data item to the text fragment from which it is derived, formulated as:

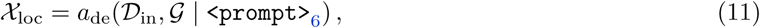

where *a*_de_ denotes the data extraction agent, D_in_ denotes the final included-study set, G denotes the predefined analysis groups,

<PROMPT>_6_ denotes the localization prompt, and X_loc_ denotes the localized source contexts and raw data extracted for the analysis groups.

**Numerical Standardization.** In the second stage, *a*_de_ standardizes the extracted raw data into the format required by the statistical synthesis engine. When necessary, it performs unit normalization and parameter conversion, such as converting standard errors to standard deviations. The final output of this module is standardized numerical data for each group that can be directly used for statistical synthesis, formulated as:

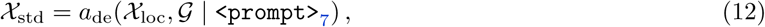

where

<PROMPT>_7_ denotes the standardization prompt, and X_std_ denotes the standardized study-level numerical evidence used for downstream evidence synthesis.

#### 4.1.6 Risk-of-Bias Assessment Agent

We formulate RoB assessment as judging the risk of bias across five domains for each included study. The RoB assessment module is defined as

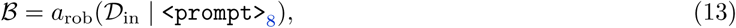

where *a*_rob_ denotes the RoB assessment agent,

<PROMPT>_8_ denotes its prompt. It takes the full texts of studies in the included-study set D_in_ as input and outputs B, the set of domain-level RoB judgments and their supporting textual evidence for the included studies across five bias domains: random sequence generation, allocation concealment, blinding of participants and personnel, blinding of outcome assessment, and incomplete outcome data. After obtaining B, the visualization engine further generates RoB summary plots to summarize the distribution of bias risk across studies.

To improve the reliability of methodological judgment, the RoB assessment module incorporates SR-RoB-7B, a task-specific model trained for RoB assessment. The training data are derived from the RoB-labeled training subset of MedSR-Bench. Each sample is constructed at the included-study level. The input consists of the full text of an included study and the RoB 1 tool, and the target output is a structured set of judgments over the annotated RoB domains. The task is formulated as a three-class classification problem, with labels including “High”, “Unclear”, and “Low”. SR-RoB-7B is initialized from Qwen2.5-7B and trained with Group Relative Policy Optimization [32] to generate structured outputs containing the predicted risk judgment and the supporting evidence for this judgment.

The reward function consists of two parts: a format reward and an accuracy reward. The format reward *r*_fmt_ constrains the model to generate complete and parseable structured outputs, including both the RoB judgments and supporting evidence. The accuracy reward *r*_acc_ measures whether the predicted judgment in each bias domain matches the ground-truth label. For a single sample, if the predicted label for the *d*-th bias domain is *y*^*_d_* and the ground-truth label is *y_d_*, the accuracy reward is defined as:

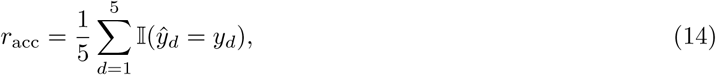

where I(·) is the indicator function, which equals 1 when the prediction matches the label and 0 otherwise. The final reward for SR-RoB-7B is:

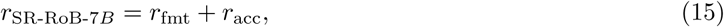

This training strategy encourages SR-RoB-7B to produce structured RoB assessment outputs while directly optimizing the correctness of domain-level risk judgments. More training details of SR-RoB-7B can be found in Appendix A.2.2.

#### 4.1.7 Evidence Synthesis Module

We formulate evidence synthesis as an analysis-group-level transformation from standardized study-level evidence to synthesized outputs. The evidence synthesis module is defined as

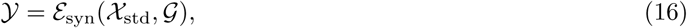

where E_syn_ denotes the statistical synthesis engine, X_std_ denotes the standardized study-level numerical evidence extracted by the data extraction module, and Y = {*y_g_* | *g* ∈ G} denotes the analysis-group-level synthesized outputs defined in Section 4.1.1.

For each analysis group, E_syn_ applies a random-effects model to combine the corresponding study-level results in X_std_ and outputs the pooled effect estimate, confidence interval, and heterogeneity statistics. The visualization engine then generates a forest plot for each analysis group using the study-level effects and the pooled effect. Based on the pooled effect and statistical results, MedSR-Copilot derives the final synthesized conclusion.

### 4.2 MedSR-Bench Construction

To systematically evaluate the capabilities of LLMs in automating the SR pipeline, we introduce **MedSR-Bench**, a benchmark curated from Cochrane reviews that supports evaluation of the full PRISMA workflow and covers 24 medical domains.

#### 4.2.1 Dataset Source and Selection Criteria

We use the **Cochrane Library** [18] as the primary data source for constructing MedSR-Bench. To ensure data quality and evaluation reliability, we select systematic reviews that satisfy the following criteria. First, each review must be published after 2023 and be freely accessible as an open-access article or through PubMed Central (PMC). Second, each review must provide the included-study list, raw data extraction results, and RoB assessment results. Third, the primary studies included in each review must be indexed in PubMed, enabling reproducible retrieval and screening evaluation.

For each eligible review, we reconstruct the structured review protocol, including PICOS information, eligibility criteria, and predefined analysis groups. When PICOS annotations are available in the original review, we directly use them; otherwise, we use Gemini 3 Flash to extract PICOS expressions from the review text and manually check the results. Gemini 3 Flash is also used to extract detailed inclusion and exclusion criteria for full-text screening.

The included-study list reported in each source review is used to construct labels for literature retrieval and literature screening. The manually checked extraction tables are used to construct ground-truth labels for data extraction and evidence synthesis. The RoB judgments reported in the source reviews are aligned with the corresponding included studies and used as labels for RoB assessment.

#### 4.2.2 Benchmark Statistics

MedSR-Bench comprises 100 systematic reviews spanning 24 distinct medical domains. The distribution of SRs across different medical domains is shown in Figure 5.

**Figure 5.**
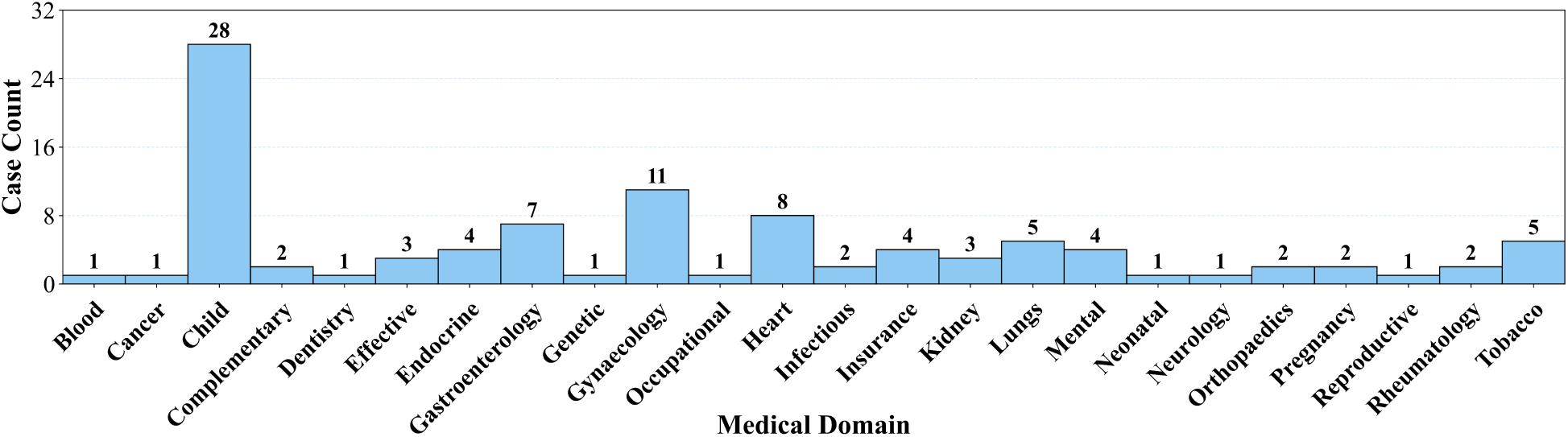
Distribution of 100 systematic reviews in MedSR-Bench across 24 medical domains.

For literature retrieval and literature screening, the benchmark samples are derived from the included-study list of each review topic. Across the 100 systematic reviews in MedSR-Bench, these lists contain 1,297 unique Randomized Controlled Trials (RCTs), which serve as the ground-truth included studies.

Data extraction and final evidence synthesis are conducted at analysis-group level. Because a single review topic can involve multiple analysis groups defined by different combinations of interventions, comparisons, and outcomes, these reviews further yield 2,144 analysis groups. For data extraction, each sample is constructed as a group-study pair, because outcome data need to be extracted separately for each included study under each predefined analysis group. This results in 7,570 data-extraction samples. For evidence synthesis, each sample is constructed at the analysis-group level, and only analysis groups with at least two included studies are used for synthesis. This results in 1,908 evidence-synthesis conclusion samples.

For RoB assessment, MedSR-Bench contains 1,204 RoB-labeled samples, which are split into 955 training samples for SR-RoB-7B and 249 test samples for evaluating MedSR-Copilot and baseline methods.

### 4.3 Evaluation Setup

**Test set construction.** For evaluations requiring full-text processing, we first construct an eligible subset of MedSR-Bench by restricting candidate reviews to those with no more than 5,000 retrieved records and more than 50% full-text availability. From this pool, we independently sample 25-review subsets for literature screening, data extraction, RoB assessment, and end-to-end evaluation. This design keeps the evaluation computationally feasible and provides a controlled basis for full-text-level tasks.

For retrieval, we use the complete sets of TrialReviewBench, TrialPanorama, and MedSR-Bench, containing 25, 100, and 100 reviews, respectively. Screening is evaluated on all 25 TrialReviewBench reviews, a 25-review TrialPanorama subset sampled using the same criteria, and the 25-review screening subset of MedSR-Bench. For data extraction, each sample is defined as an analysis-group–study pair, resulting in 412 samples from Yun Bench and 3,610 samples from the 25-review extraction subset of MedSR-Bench. For RoB assessment, each RCT is treated as one sample, resulting in 127 test samples from RoBBR and 249 test samples from MedSR-Bench. For end-to-end evaluation, the 25-review subset yields 316 end-to-end conclusion samples.

**Task protocols and baseline adaptation.** For end-to-end conclusion evaluation, all methods start from the same research protocol and follow the same staged workflow, where the output of each preceding task is passed to the next task. The extracted standardized data from all methods are then processed by the same statistical synthesis engine to generate synthesized conclusions. RoB assessment is not executed as an intermediate step in end-to-end conclusion evaluation, because its outputs are not the required inputs to the statistical synthesis engine.

For retrieval, automated systematic-review systems generate search queries using their original workflows. DeepRetrieval is minimally adapted to accept PICOS information for query rewriting, while the standalone LLM baselines use the same prompt template to directly generate search queries.

For screening, each review is assigned a fixed candidate pool of 2,000 studies, containing all ground-truth included studies and additional non-ground-truth studies sampled from the retrieved candidate set. MedSR-Copilot and Manalyzer perform full-text-level screening, using titles and abstracts as fallback when full texts are unavailable, whereas the remaining baselines use title and abstract information only.

For data extraction, standalone LLM baselines directly output standardized data, whereas TrialMindSLR and MedSR-Copilot both use multi-step extraction. However, TrialMindSLR is not explicitly designed to first localize raw evidence for multiple predefined analysis groups; instead, it standardizes from cohort-level result summaries, while MedSR-Copilot standardizes from raw evidence of each analysis group.

For RoB assessment, standalone LLM baselines and SR-RoB-7B directly output judgments for the five RoB domains, while TrialMindSLR is adapted by treating the RoB tool and domain definitions as extraction descriptions.

MedSR-Copilot uses Gemini 3.1 Flash-Lite for the end-to-end evaluation and for all subtask evaluations except the RoB assessment subtask, where SR-RoB-7B is used as the RoB assessment component.

**Evaluation metrics.** For literature retrieval, we use Recall@3000 and Recall@5000. For each review, we compare the retrieved records with the ground-truth included-study list and calculate the proportion of included studies captured within the top 3,000 and top 5,000 retrieved results, respectively.

For literature screening, we use F1 score as the primary metric, with Recall and Precision reported as complementary metrics. Recall and Precision are computed by comparing the final included-study list with the ground-truth included-study list.

For data extraction, we report Group Recall and Data-item Accuracy. Group recall measures the proportion of target analysis groups for which the method successfully identifies corresponding extractable data, whereas data-item accuracy measures the proportion of standardized numerical values that match the ground-truth labels.

For RoB assessment, we formulate judgment for each domain as a three-class classification problem, where the method predicts whether the risk level of each RoB domain is high, unclear, or low. We evaluate RoB domains with annotated labels for each RCT. The Accuracy of each RCT sample is calculated as the average judgment accuracy across its five RoB domains, and the final RoB accuracy is reported as the average across all RCT samples.

For end-to-end evidence synthesis, we formulate final conclusion evaluation as a three-class classification task: favoring the intervention, favoring the control, or showing no significant effect. We report Accuracy, defined as the proportion of analysis groups for which the predicted conclusion category matches the corresponding human-derived SR conclusion.

## Supporting information

All supplementary files of the main paper

## Data Availability

https://huggingface.co/datasets/halfmorepiece/MedSR-Bench

https://huggingface.co/datasets/RoBBR-Benchmark/RoBBR/tree/main

https://huggingface.co/datasets/TrialPanorama/Dataset

## Footnotes

1 PRISMA stands for “Preferred Reporting Items for Systematic reviews and Meta-Analyses” https://www.prisma-statement.org/.

2 Population, Intervention, Comparison, Outcome, and Study Design

## Acknowledgments

This work was supported by the Shanghai Municipal Special Program for Basic Research on General Al Foundation Models (Grant No. 2025SHZDZX026D12).

## 5 Author Contributions

All listed authors clearly meet the ICMJE 4 criteria. F.W., X.Z. and C.W. are the corresponding authors. Specifically, H.H., Q.Z., P.Q., W.Z., Y.Z, W.X, F.W., X.Z. and C.W. all make contributions to the conception or design of the work, and H.H. further performs acquisition, analysis, or interpretation of data for the work. In writing, H.H. drafts the work. Q.Z., P.Q., W.Z., Y.Z, W.X, F.W., X.Z. and C.W. review it critically for important intellectual content. All authors approve of the version to be published and agree to be accountable for all aspects of the work to ensure that questions related to the accuracy or integrity of any part of the work are appropriately investigated and resolved.

## 6 Code and Dataset Availability

The source code developed for this study is publicly available at https://github.com/MAGIC-AI4Med/MedSR-Copilot. The MedSR-Bench dataset is available at https://huggingface.co/datasets/halfmorepiece/MedSR-Bench.

## References

[1] David L Sackett. Evidence-based medicine. In Seminars in perinatology, volume 21, pages 3–5. Elsevier, 1997.

[2] Andy P Field and Raphael Gillett. How to do a meta-analysis. British Journal of Mathematical and Statistical Psychology, 63(3):665–694, 2010.

[3] John Concato, Nirav Shah, and Ralph I Horwitz. Randomized, controlled trials, observational studies, and the hierarchy of research designs. In Research ethics, pages 207–212. Routledge, 2017.

[4] Ursula Griebler, Dominic Ledinger, Irma Klerings, Stefan Schandelmaier, Andreea Dobrescu, Mieke Deschodt, Stuart McLennan, Lars G Hemkens, Barbara Nussbaumer-Streit, and Matthias Briel. Identifying prior evidence for new trials (reveal): guidance for clinical researchers. bmj, 392, 2026.

[5] Jacqueline Chandler, Miranda Cumpston, Tianjing Li, Matthew J Page, VJHW Welch, et al. Cochrane handbook for systematic reviews of interventions. Hoboken: Wiley, 4(1002):14651858, 2019.

[6] Matthew J Page, Joanne E McKenzie, Patrick M Bossuyt, Isabelle Boutron, Tammy C Hoffmann, Cynthia D Mulrow, Larissa Shamseer, Jennifer M Tetzlaff, Elie A Akl, Sue E Brennan, et al. The prisma 2020 statement: an updated guideline for reporting systematic reviews. bmj, 372, 2021.

[7] Rohit Borah, Andrew W Brown, Patrice L Capers, and Kathryn A Kaiser. Analysis of the time and workers needed to conduct systematic reviews of medical interventions using data from the prospero registry. BMJ open, 7(2):e012545, 2017.

[8] Zhen Wang, Tarek Nayfeh, Jennifer Tetzlaff, Peter O’Blenis, and Mohammad Hassan Murad. Error rates of human reviewers during abstract screening in systematic reviews. PloS one, 15(1):e0227742, 2020.

[9] Julian Elliott, Rebecca Lawrence, Jan C Minx, Olufemi T Oladapo, Philippe Ravaud, Britta Tendal Jeppesen, James Thomas, Tari Turner, Per Olav Vandvik, and Jeremy M Grimshaw. Decision makers need constantly updated evidence synthesis. Nature, 600(7889):383–385, 2021.

[10] Chris Lu, Cong Lu, Robert Tjarko Lange, Jakob Foerster, Jeff Clune, and David Ha. The ai scientist: Towards fully automated open-ended scientific discovery. arXiv preprint arXiv:2408.06292, 2024.

[11] Siddhant Jain, Asheesh Kumar, Trinita Roy, Kartik Shinde, Goutham Vignesh, and Rohan Tondulkar. Scispace literature review: harnessing ai for effortless scientific discovery. In European Conference on Information Retrieval, pages 256–260. Springer, 2024.

[12] Akari Asai, Jacqueline He, Rulin Shao, Weijia Shi, Amanpreet Singh, Joseph Chee Chang, Kyle Lo, Luca Soldaini, Sergey Feldman, Mike D’Arcy, et al. Synthesizing scientific literature with retrieval-augmented language models. Nature, pages 1–7, 2026.

[13] Ziru Chen, Shijie Chen, Yuting Ning, Qianheng Zhang, Boshi Wang, Botao Yu, Yifei Li, Zeyi Liao, Chen Wei, Zitong Lu, et al. Scienceagentbench: Toward rigorous assessment of language agents for data-driven scientific discovery. In *International Conference on Learning Representations*, volume 2025, pages 96934–96990, 2025.

[14] Zifeng Wang, Lang Cao, Benjamin Danek, Qiao Jin, Zhiyong Lu, and Jimeng Sun. Accelerating clinical evidence synthesis with large language models. npj Digital Medicine, 8(1):509, 2025.

[15] Zifeng Wang, Qiao Jin, Jiacheng Lin, Junyi Gao, Jathurshan Pradeepkumar, Pengcheng Jiang, Benjamin Danek, Zhiyong Lu, and Jimeng Sun. Trialpanorama: Database and benchmark for systematic review and design of clinical trials. arXiv e-prints, pages arXiv–2505, 2025.

[16] HS Yun, D Pogrebitskiy, IJ Marshall, and BC Wallace. Automatically extracting numerical results from randomized controlled trials with large language models. arXiv preprint arXiv:2410.03230, 2024.

[17] Jianyou Wang, Weili Cao, Longtian Bao, Youze Zheng, Gil Pasternak, Kaicheng Wang, Xiaoyue Wang, Ramamohan Paturi, and Leon Bergen. Measuring risk of bias in biomedical reports: The robbr benchmark. In Proceedings of the 2025 Conference on Empirical Methods in Natural Language Processing, pages 3220–3248, 2025.

[18] Cochrane Collaboration. Cochrane library, 2026. Accessed: 2026–04-11.

[19] Pengcheng Jiang, Jiacheng Lin, Lang Cao, Runchu Tian, SeongKu Kang, Zifeng Wang, Jimeng Sun, and Jiawei Han. Deepretrieval: Hacking real search engines and retrievers with large language models via reinforcement learning. *arXiv preprint arXiv:2503.00223*, 2025.

[20] Wanghan Xu, Wenlong Zhang, Fenghua Ling, Ben Fei, Yusong Hu, Runmin Ma, Bo Zhang, Fangxuan Ren, Jintai Lin, Wanli Ouyang, et al. Manalyzer: End-to-end automated meta-analysis with multi-agent system. arXiv preprint arXiv:2505*.20310*, 2025.

[21] Yiqun Chen, Qi Liu, Yi Zhang, Weiwei Sun, Xinyu Ma, Wei Yang, Daiting Shi, Jiaxin Mao, and Dawei Yin. Tourrank: Utilizing large language models for documents ranking with a tournament-inspired strategy. In Proceedings of the ACM on Web Conference 2025, pages 1638–1652, 2025.

[22] Julian PT Higgins, Douglas G Altman, Peter C Gøtzsche, Peter Jüni, David Moher, Andrew D Oxman, Jelena Savović, Kenneth F Schulz, Laura Weeks, and Jonathan AC Sterne. The cochrane collaboration’s tool for assessing risk of bias in randomised trials. bmj, 343, 2011.

[23] Iain J Marshall and Byron C Wallace. Toward systematic review automation: a practical guide to using machine learning tools in research synthesis. Systematic reviews, 8(1):163, 2019.

[24] Honghao Lai, Jiayi Liu, Chunyang Bai, Hui Liu, Bei Pan, Xufei Luo, Liangying Hou, Weilong Zhao, Danni Xia, Jinhui Tian, et al. Language models for data extraction and risk of bias assessment in complementary medicine. Npj Digital Medicine, 8(1):74, 2025.

[25] Lingbo Li, Anuradha Mathrani, and Teo Susnjak. Transforming evidence synthesis: a systematic review of the evolution of automated meta-analysis in the age of ai. arXiv preprint arXiv:2504.20113, 2025.

[26] Elsevier. Embase, 2026. Biomedical and pharmacological database, 1947–present.

[27] Cochrane Collaboration. Cochrane central register of controlled trials (central), 2026. Database of randomized and quasi-randomized controlled trials.

[28] Eric Sayers and David Wheeler. Building customized data pipelines using the entrez programming utilities (eUtils). NCBI, 2004.

[29] Henry J Lowe and G Octo Barnett. Understanding and using the medical subject headings (mesh) vocabulary to perform literature searches. Jama, 271(14):1103–1108, 1994.

[30] Abdelrahman Abdallah Bhawna Piryani Jamshid Mozafari and Mohammed Ali Adam Jatowt. How good are llm-based rerankers? an empirical analysis of state-of-the-art reranking models. 2025.

[31] Gongbo Zhang, Qiao Jin, Denis Jered McInerney, Yong Chen, Fei Wang, Curtis L Cole, Qian Yang, Yanshan Wang, Bradley A Malin, Mor Peleg, et al. Leveraging generative ai for clinical evidence synthesis needs to ensure trustworthiness. Journal of biomedical informatics, 153:104640, 2024.

[32] Zhihong Shao, Peiyi Wang, Qihao Zhu, Runxin Xu, Junxiao Song, Xiao Bi, Haowei Zhang, Mingchuan Zhang, Y. K. Li, Y. Wu, and Daya Guo. Deepseekmath: Pushing the limits of mathematical reasoning in open language models. arXiv preprint arXiv:2402*.03300*, 2024.

