## Supplementary material for "A PRISMA-Aligned Agentic Framework for Medical Systematic Reviews and Evidence Synthesis": All supplementary files of the main paper

### A Supplementary

#### A.1 Supplementary Results

##### A.1.1 Results of Human-AI Collaboration Experiment in Literature Screening and Data Extraction

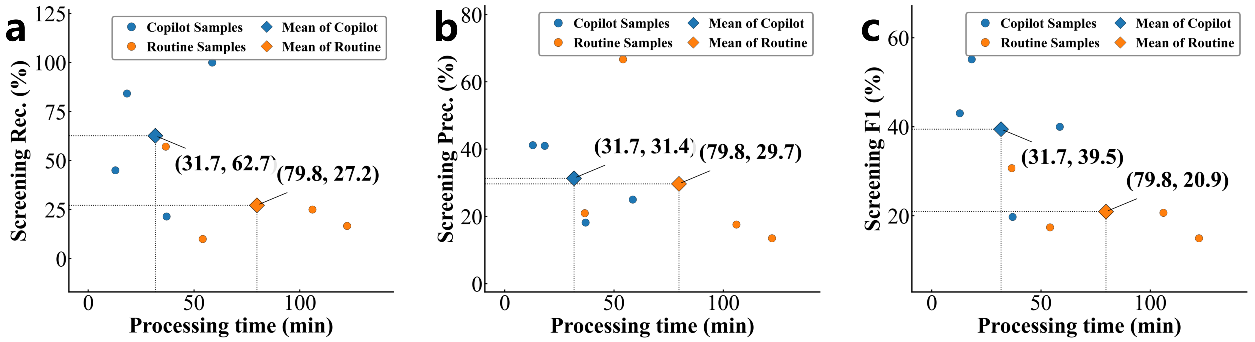


**Supplementary Figure 1** | Human–AI collaboration results on literature screening. **a**, Screening recall and processing time. **b**, Screening precision and processing time. **c**, Screening F1 score and processing time. Each panel compares the routine-practice SR condition with the MedSR-Copilot-augmented condition.

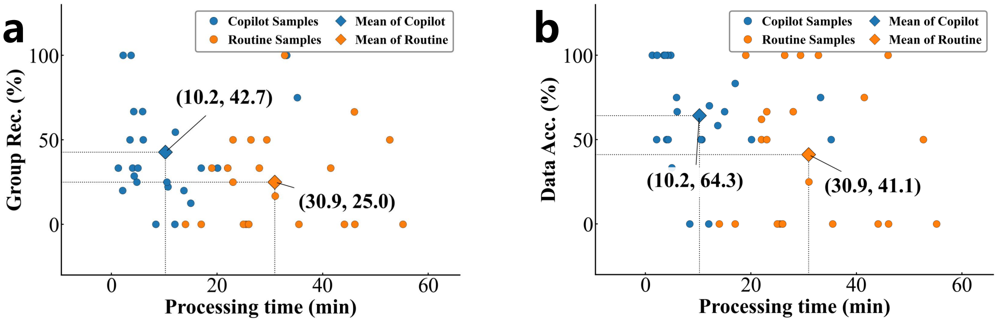


**Supplementary Figure 2** | Human–AI collaboration results on data extraction. **a**, Analysis-group recall and processing time. **b**, Data-item accuracy and processing time. Each panel compares the routine-practice SR condition with the MedSR-Copilot-augmented condition.

Supplementary Figure 1 and Supplementary Figure 2 further summarize the time–performance trade-offs for the screening and extraction subtasks. In screening, the average processing time decreases from 79.8 to 31.7 minutes, while the F1 score increases from 20.9% to 39.5%, screening recall increases from 27.2% to 62.7%, and screening precision increases from 29.7% to 31.4%. In data extraction, the average processing time decreases from 30.9 to 10.2 minutes, while analysis-group recall improves from 25.0% to 42.7% and data-item accuracy improves from 41.1% to 64.3%. These results show that MedSR-Copilot not only accelerates the review process for participants but also improves the quality of key intermediate outputs passed to downstream evidence synthesis.

##### A.1.2 Human–AI Collaboration Usability Evaluation

To assess participants' perceptions of the MedSR-Copilot platform, we provide each participant with a structured questionnaire covering five dimensions: Ease of Use, Workflow Alignment, Output Quality, Transparency, and Visualization Quality. Each dimension contains four sub-questions, and each sub-question is scored on a 1–5 Likert scale. A score of 1 indicates strong disagreement or very poor experience, 2 indicates disagreement or poor experience, 3 indicates a neutral or basically acceptable experience, 4 indicates agreement or good experience, and 5 indicates strong agreement or very good experience. The score of each dimension is calculated as the average score across its sub-questions, and the overall usability score can be calculated as the average across the five dimensions.

The five dimensions are defined as follows. Ease of Use evaluates whether the interface is intuitive, easy to operate, and has a low learning cost. Workflow Alignment evaluates whether the platform follows the standard workflow of systematic reviews and meta-analyses and naturally supports standard systematic-review practice. Output Quality evaluates the reliability and usability of the generated screening, extraction, RoB assessment, and synthesis outputs. Transparency evaluates whether intermediate results, evidence traces, and reasoning bases are visible and traceable. Visualization Quality evaluates the clarity, standardization, and practical usefulness of generated visual outputs such as PRISMA flow diagrams, forest plots, and RoB plots.

As shown in Supplementary Figure 3, participants give consistently positive ratings across all five dimensions. The highest average score is observed for Ease of Use, reaching 4.7, suggesting that participants can operate the platform with a low learning cost. Visualization Quality also receives a high average score of 4.3, indicating that the generated diagrams are clear and useful for communication and result inspection. Workflow Alignment reaches 4.2, showing that the platform is broadly consistent with standard systematic-review practice. Output Quality and Transparency both reach 4.1, suggesting that participants regard the generated outputs as useful for human review and find most intermediate results traceable. Overall, the usability results indicate that MedSR-Copilot is not only effective in task performance but also acceptable as a researcher-facing collaborative platform.

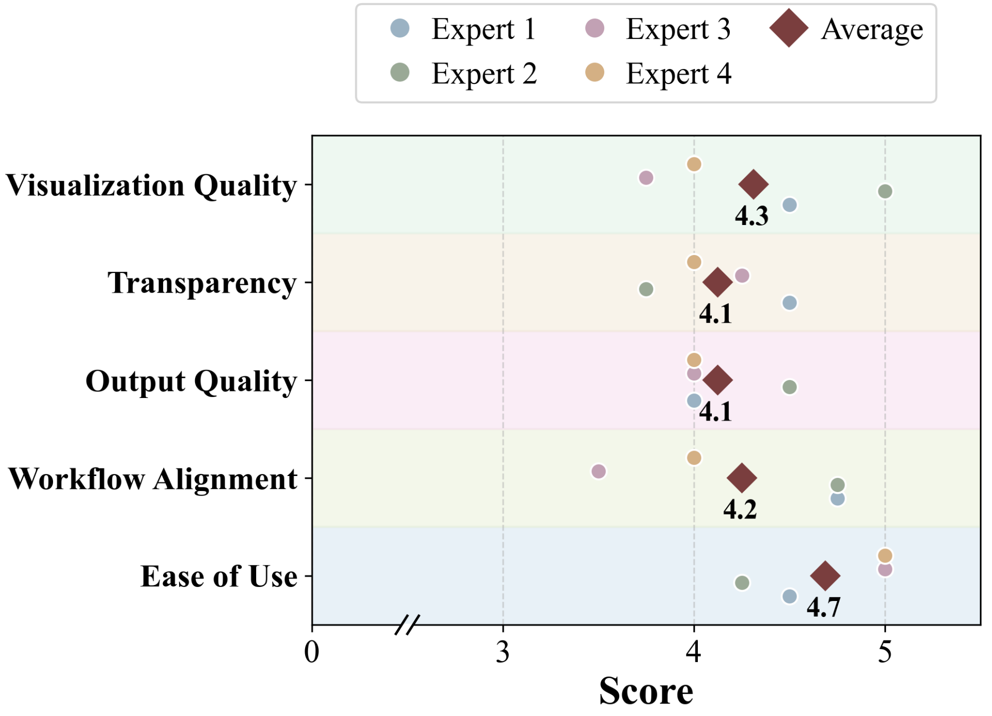


**Supplementary Figure 3** | Usability evaluation of the MedSR-Copilot platform by four participants. Five dimensions are scored on a 1–5 scale, including Ease of Use, Workflow Alignment, Output Quality, Transparency, and Visualization Quality. Circles denote individual expert ratings, and diamonds denote average scores.

##### A.1.3 End-to-End Ablation Variant Definitions

For the end-to-end ablation study, each variant follows the same evaluation protocol as the full MedSR-Copilot system and only replaces the target component under examination. For the variant without tournament-style reranking, we directly use the studies that pass full-text screening as the final included studies. For the variant without Review-RAG, we set the reference systematic-review content to empty during query generation. For the variant without two-stage data extraction, the model directly outputs standardized meta-analytic data items without first locating the supporting source context.

##### A.1.4 Ablation Study on Review-RAG Module

To further examine the contribution of Review-RAG, we conduct ablation experiments on all three retrieval benchmarks. As shown in Supplementary Figure 4, removing Review-RAG consistently reduces retrieval recall. Compared with the non-RAG variant, the full MedSR-Copilot improves Recall@3000 by 2.6, 4.2, and 3.7 percentage points on TrialReviewBench, TrialPanorama, and MedSR-Bench, respectively. The same trend is observed for Recall@5000, where MedSR-Copilot improves performance by 3.0, 3.9, and 4.4 percentage points, respectively. These results indicate that retrieving relevant systematic reviews as domain references helps generate broader and more precise PubMed queries.

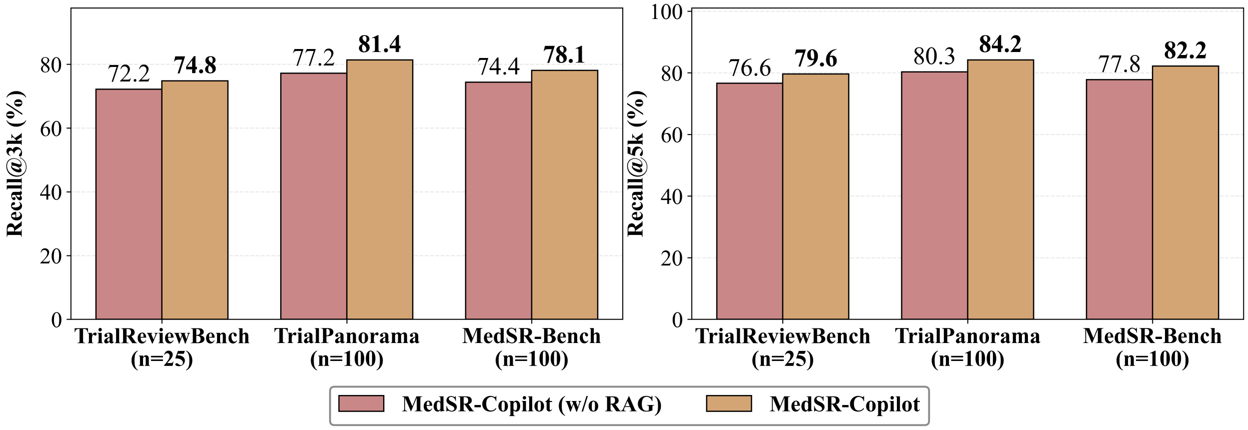


**Supplementary Figure 4** | Ablation study of Review-RAG in the retrieval module. Recall@3000 and Recall@5000 are reported as percentages on TrialReviewBench, TrialPanorama, and MedSR-Bench.

##### A.1.5 Ablation Study of Full-text Reranking in Literature Screening

We further conduct an ablation study to examine the effect of full-text reranking in the literature screening module. As shown in Supplementary Figure 5, full-text reranking consistently improves the F1 score across all three benchmarks.

On TrialReviewBench, adding reranking increases F1 from 19.5% to 46.3%, corresponding to a gain of 26.8 percentage points. Precision increases from 34.2% to 43.4%, while Recall decreases from 64.6% to 56.2%. On TrialPanorama, F1 increases from 24.7% to 43.9%, a gain of 19.2 percentage points, with Precision increasing from 38.4% to 41.3% and Recall decreasing slightly from 66.4% to 63.3%. On MedSR-Bench, F1 increases from 44.7% to 51.0%, a gain of 6.3 percentage points, with Precision increasing from 43.9% to 47.2% and Recall decreasing from 80.1% to 62.1%.

These results show that full-text reranking enhances the discriminative ability of the screening module by prioritizing studies that are more consistent with the review question and detailed eligibility criteria. Although reranking leads to some loss in recall, the consistent improvements in F1 score indicate a better overall balance between retaining eligible studies and excluding false positives. The accompanying precision gains mean that fewer non-ground-truth studies are passed to downstream stages. This is particularly important for evidence synthesis, because reducing irrelevant or weakly matched studies helps alleviate the interference of noisy evidence in data extraction and final conclusion generation.

##### A.1.6 Risk-of-Bias Assessment Results Across 5 RoB Domains

We further analyze RoB assessment performance across the five domains in RoB 1 tool, as shown in Supplementary Figure 6. On RoBBR, all methods achieve their highest performance on random sequence generation, with Gemini 3 Flash and GPT-5.4-Mini reaching 82.1%, DeepSeek-v4-flash 80.5%, MedSR-Copilot 79.7%, and TrialMindSLR 75.6%. In contrast, incomplete outcome data is more challenging for most baselines,

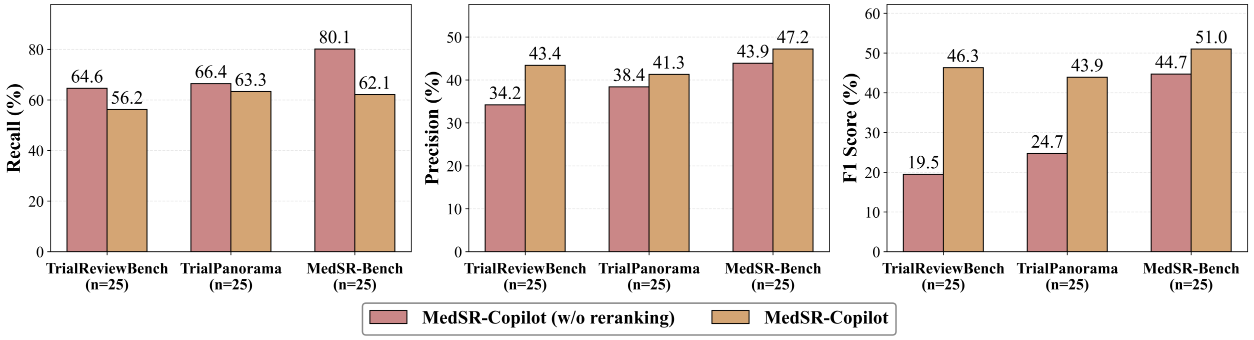


**Supplementary Figure 5** | Ablation study of full-text reranking in literature screening. Recall, Precision and F1 score are reported as percentages on TrialReviewBench, TrialPanorama, and MedSR-Bench, comparing MedSR-Copilot with and without the reranking module.

where DeepSeek-v4-flash, Gemini 3 Flash, GPT-5.4-Mini, and TrialMindSLR obtain 46.8%, 40.5%, 37.3%, and 50.0%, respectively, while MedSR-Copilot achieves 69.8%. MedSR-Copilot also obtains the highest accuracy on blinding of participants and personnel and incomplete outcome data, suggesting stronger performance in domains requiring detailed methodological judgment.

On the RoB test subset of MedSR-Bench, MedSR-Copilot achieves the highest micro-average accuracy of 70.1%, compared with 68.0% for DeepSeek-v4-flash, 64.5% for Gemini 3 Flash, 63.6% for GPT-5.4-Mini, and 56.6% for TrialMindSLR. Domain-level results show that MedSR-Copilot performs best on random sequence generation and incomplete outcome data, reaching 77.7% and 77.1%, respectively. Across the other three domains, DeepSeek-v4-Flash achieved the best performance in allocation concealment with 70.1%, Gemini 3 Flash in blinding of participants and personnel with 74.5%, and GPT-5.4-Mini in blinding of outcome assessment with 66.4%. Overall, MedSR-Copilot shows more balanced performance across RoB domains and a clear advantage on incomplete outcome data.

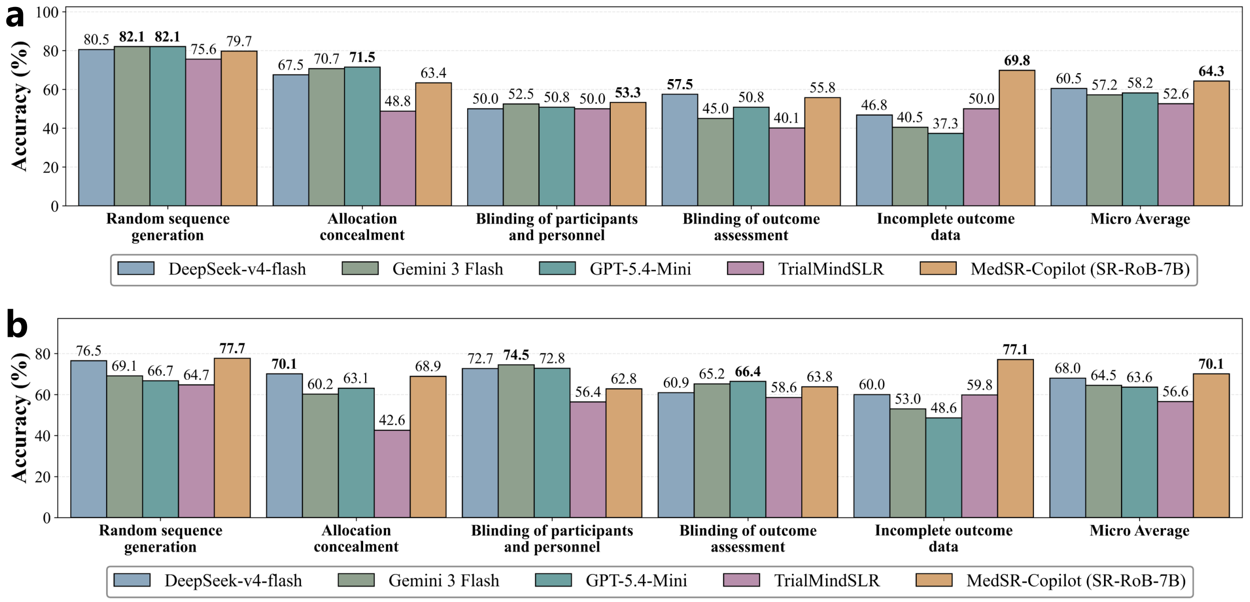


**Supplementary Figure 6** | Risk-of-Bias assessment performance across five RoB domains. The comparison includes DeepSeek-v4-flash, Gemini 3 Flash, GPT-5.4-Mini, TrialMindSLR, and MedSR-Copilot on RoBBR and MedSR-Bench. **a**, RoB judgment accuracy on RoBBR, which includes 127 studies and each study with a single RoB domain to be evaluated. **b**, RoB judgment accuracy on the test subset of MedSR-Bench, which includes 249 studies and each study with multiple RoB domains to be evaluated.

#### A.2 Details of Methods

##### A.2.1 Tournament-inspired Full-text Reranking

The full-text reranking module is designed to improve the discriminative ability of the screening stage after preliminary full-text eligibility assessment. Standard pointwise screening only determines whether each study satisfies the predefined criteria independently, but it is often insufficient when multiple candidate studies share similar populations, interventions, outcomes, and trial designs. In this setting, several studies may pass the full-text screening stage while only a subset of them is most consistent with the target systematic review. Therefore, we introduce a tournament-inspired reranking procedure to perform relative comparison among candidate studies.

Given the set of preliminarily included full-text candidates, the reranking module takes the full PICOS information, detailed inclusion and exclusion criteria, and the full texts of candidate studies as input. The model is required to compare studies within a small group and select the candidates that best match the target review question. This design avoids directly applying listwise ranking to a large number of full texts, which is constrained by the context length of LLMs, and also avoids exhaustive pairwise comparison, which is computationally expensive for full-text inputs.

Specifically, MedSR-Copilot runs $R = 8$ independent tournament instances in parallel. For each tournament, the candidate studies are first randomly shuffled and then partitioned into groups of $g = 5$ studies. Within each group, the model compares the five full-text studies according to the target population, intervention, comparison, outcome, study design, and detailed eligibility criteria, and selects the top $m = 2$ studies to advance to the next round. Each time a study advances, it receives one advancement point. The same grouping, comparison, and promotion process is repeated until the final top two studies are obtained in that tournament. To reduce the influence of a single random grouping, the initial candidate order is independently shuffled across the eight tournaments.

After all tournaments are completed, the final reranking score of a study is defined as the cumulative number of advancement points it receives across all tournament instances:

$$s_i = \sum_{r=1}^{R} a_{i,r},$$

where $a_{i,r}$ denotes the number of advancement points obtained by study $i$ in the $r$-th tournament. Candidate studies are then sorted in descending order according to $s_i$. The top $k$ studies are retained as the final included studies for downstream data extraction and evidence synthesis. In our screening experiments, we set the reranking cutoff to $k = 20$ and retain the top-20 ranked studies as the final candidates. If the number of preliminarily included full-text candidates does not exceed $k$, the reranking step is skipped and all candidates are retained.

This tournament-inspired design improves the screening module in two ways. First, it converts difficult full-text screening from isolated yes/no decisions into local relative comparisons, allowing the model to better distinguish studies that are highly similar but differ in their alignment with detailed eligibility criteria. Second, repeated randomized tournaments make the final ranking more robust than a single group comparison, because each study can be evaluated under multiple local comparison contexts. The resulting ranked list is then used as the final inclusion list passed to the data extraction module.

##### A.2.2 Training of SR-RoB-7B

SR-RoB-7B is a task-specific model trained for Risk-of-Bias assessment under the Cochrane RoB 1 tool. To keep the training setting consistent with the RoB evaluation protocol in MedSR-Bench, we use the RoB-labeled samples from MedSR-Bench and split them into a training subset of 955 samples and a held-out test subset of 249 samples. The test subset is used only for evaluating MedSR-Copilot, SR-RoB-7B, and all baseline methods. For model training, the 955 training samples are further divided into 855 training samples and 100 validation samples with a fixed random seed of 42.

Each sample is constructed at the included-study level. The input consists of the full text of an included study and the RoB 1 tool. The target output is a structured domain-level judgment over five bias domains, including random sequence generation, allocation concealment, blinding of participants and personnel, blinding of outcome assessment, and incomplete outcome data. Each domain is labeled as one of three risk levels: "High", "Unclear", or "Low". During preprocessing, RoB labels are normalized to these three categories, and samples without valid RoB annotations are removed.

SR-RoB-7B is initialized from Qwen2.5-7B and optimized with Group Relative Policy Optimization (GRPO) [32] using the verl framework. The reward function is rule-based and combines output-format validation with domain-level label accuracy. The format reward checks whether the response is a parseable JSON object that contains valid judgments for all five RoB domains. Invalid outputs receive a reward of $-1$. For valid outputs, we calculate the accuracy reward by comparing the predicted judgment with the ground-truth label over the available annotated domains:

$$r_{\text{acc}}(i) = \frac{1}{|\mathcal{D}_i|} \sum_{d \in \mathcal{D}_i} \mathbb{I}\left(\hat{y}_{i,d} = y_{i,d}\right),$$

where $\mathcal{D}_i$ denotes the set of annotated RoB domains for sample $i$. The final reward is defined as:

$$r_{\text{RoB}}(i) = \begin{cases} -1, & \text{if the output format is invalid,} \\ 1 + r_{\text{acc}}(i), & \text{if the output format is valid.} \end{cases}$$

This reward design encourages the model to produce structured outputs while directly optimizing the correctness of final RoB risk-level judgments.

We train SR-RoB-7B on 8 NVIDIA A100 GPUs with 80 GB memory. The total number of training epochs is 15. The training batch size is 8, and each prompt samples $n = 8$ responses for GRPO, resulting in a PPO mini-batch size of 64. The PPO micro-batch size per GPU is 2. The maximum prompt length is 15,000 tokens, and the maximum response length is 800 tokens. The learning rate is $5 \times 10^{-6}$, with a warmup ratio of 0.02, a minimum learning-rate ratio of 0.1, and a cosine warmup schedule. KL regularization is applied to the actor loss with a coefficient of 0.001. We use vLLM for rollout generation and enable gradient checkpointing, padding removal, and FSDP parameter offloading for memory-efficient training. Validation is performed every 10 steps, and the checkpoint with the best performance on validation set is saved.

#### A.3 Platform for MedSR-Copilot

To support practical use of MedSR-Copilot, we implement an interactive web platform that allows users to submit a systematic-review protocol and inspect the outputs generated along the PRISMA-aligned workflow. As shown in Supplementary Figure 7, the platform first accepts a natural-language research protocol as input. The system then parses the protocol into structured review elements, including PICOS information, detailed eligibility criteria, and analysis-group definitions. Users can directly edit or supplement these fields before executing the downstream workflow, which allows domain experts to refine the review question and align the system input with the intended clinical scope.

After the protocol is confirmed, MedSR-Copilot sequentially performs literature retrieval, hierarchical literature screening, data extraction, Risk-of-Bias assessment, and evidence synthesis. The platform is designed to expose not only the final synthesis results but also the intermediate records produced by each module. Specifically, it provides the PubMed retrieval records, screening decisions and exclusion reasons, the PRISMA flow diagram, the final included-study list, structured extraction tables, RoB judgments, forest plots, and synthesized conclusions for each analysis group. This design enables users to trace how the final conclusion is derived from the original research protocol and candidate studies, rather than treating the system as a black-box generator.

The platform is intended to function as a human–AI collaborative interface for systematic reviews. Users can verify the structured protocol, check the included studies, examine extracted numerical evidence, and review the generated synthesis results. By organizing these outputs within a single interface, the platform makes the MedSR-Copilot workflow more transparent and facilitates expert review of key intermediate artifacts.

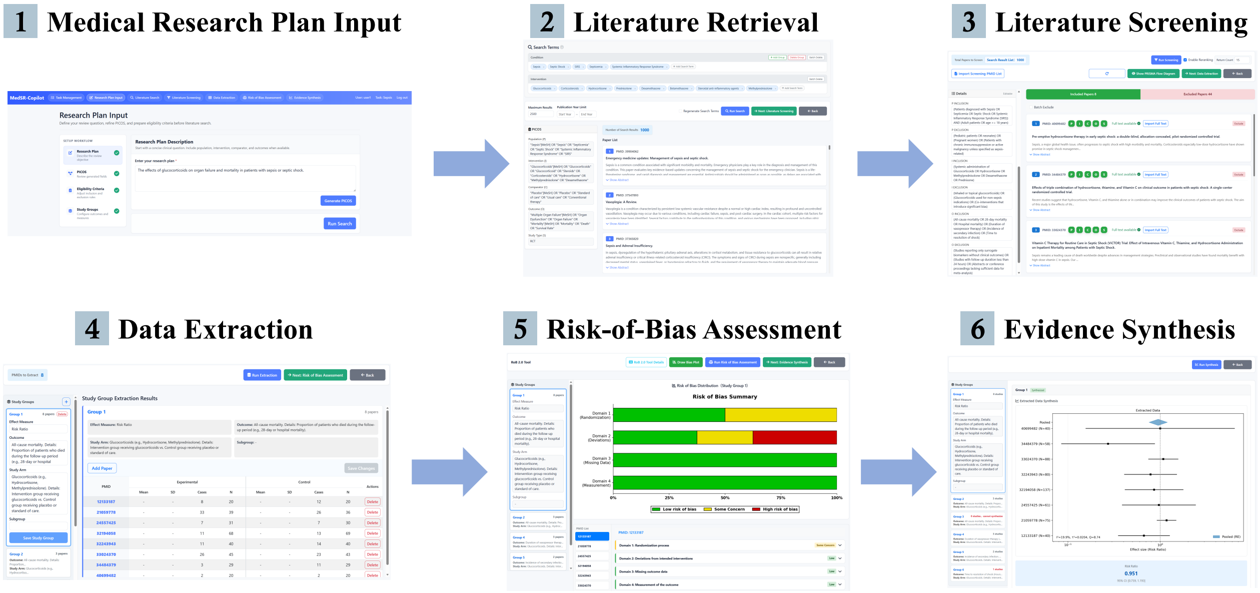


**Supplementary Figure 7** | Platform interface of MedSR-Copilot. The platform supports protocol submission, structured review-element generation, and inspection of intermediate records and final evidence-synthesis outputs, including retrieval records, screening results, extracted data, Risk-of-Bias judgments, forest plots, and synthesized conclusions.

##### A.4 Prompt Collection

**Prompt 1. Prompt for `literature retrieval` subagent with reference systematic reviews and PI as input**

**Background**

You are a clinical specialist. You are conducting a clinical meta-analysis.

The research is defined by the following P and I terms:

- P (Patient, Problem or Population): {P}
- I (Intervention): {I}

**Reference Systematic Reviews / Meta-Analyses**

You've gathered the following **Systematic Reviews and Meta-Analyses** as reference literature. These reviews have conducted comprehensive literature searches on topics related (but not necessarily identical) to your research question:

{Reference}

**Instruction for using the reference systematic reviews:**

1. **Analyze their P and I scope**: Carefully examine how each systematic review defined their research population (P) and intervention (I). Identify the key themes and inclusion criteria they used.
2. **CRITICAL - Adapt, Don't Copy**:
   - If the reference reviews' topics are **exactly aligned** with your P and I terms above, you may reference their search strategies closely.
   - **However, if the reference reviews' topics are NOT perfectly matched with your P and I terms:**
     - **DO NOT simply copy their search strategies or analysis group definitions**
     - Use their strategies as **INSPIRATION and REFERENCE** only
3. **Priority**: Your generated search terms MUST always primarily reflect the USER's P and I terms provided above, NOT the reference reviews' topics.

**Task**

Your task is to generate the final search terms for this meta-analysis.

TWO STEP GENERATION PROCESS:

You should reason in BRIEF FREETEXT REASONING PROCESS with this order:

1. INITIAL GENERATION: simplify the target question and identify the most central disease / condition concepts from P and the most central intervention concepts from I. Use a mix of precise MeSH-aligned terminology and free-text clinical terms when helpful.

2. AUGMENTATION: expand the final terms with precise synonyms, full forms of abbreviations, and closely related free-text variants.

Generate two final lists:

1. CONDITIONS: final search terms for the population / condition / disease concepts in P.
2. TREATMENTS: final search terms for the intervention concepts in I.

**Reply format**

BRIEF FREETEXT REASONING PROCESS, NO MORE THAN 50 WORDS. NO MORE THAN 15 TERMS IN EACH LIST.

Then, your reply must end with a JSON object like this:

```
{
"CONDITIONS": ["term1", "term2", "term3"],
"TREATMENTS": ["term1", "term2", "term3"]
}
```

**Prompt 2. Prompt for `literature screening` subagent: title-level screening**

#### CONTEXT

You are a clinical specialist tasked with assessing research papers for inclusion in a meta-analysis.

#### OBJECTIVE

Evaluate whether the paper TITLE indicates that the study matches the specified Population (P) and Intervention (I) for inclusion in the meta-analysis. Provide decisions ("YES", "NO", or "UNCERTAIN") for P and I.

#### JUDGEMENT INSTRUCTION

- **YES**: Title explicitly reports the target Population or Intervention or clearly equivalent in the given context
- **UNCERTAIN**:

1. Title doesn't give information about any kind of Population or Intervention
2. Title mentions part of target P or I, but does not meet the entire requirement. For example, 'Chemotherapy and radiation therapy' in TITLE, I is "Chemotherapy"

- **NO**: The title explicitly reports a Population or Intervention that shares no consistent components with the target P or I, nor is it synonymous

#### RESEARCH FRAMEWORK

- P (Patient, Problem or Population): {P}

- I (Intervention): {I}

#### PAPER DETAILS

- Provided Paper Title: {paper_title}

#### RESPONSE FORMAT

1. ANALYSIS: you should first output a **BRIEF, LESS THAN 30 WORDS** analysis based on the source context, through identifying synonyms (e.g., 'any grade' and 'grade 1-5'), specific examples or sub-types (e.g., 'cisplatin' as a subtype of 'chemotherapy'), and hypernym (e.g., 'AE' as a hypernym of 'irAE') for P and I. For example:

   "ANALYSIS": ["From 'marijuana relieves pain in oncology patients' in TITLE, 'oncology patients' is a synonym for 'cancer patients', matches P.", ... ] // List of 2 items for P and I
2. Evaluations: provide your final decisions for P and I in order based on the ANALYSIS: "evaluations": ["YES/NO/UNCERTAIN for P", "YES/NO/UNCERTAIN for I"]

**Prompt 3. Prompt for `literature screening` subagent: abstract-level screening**

#### CONTEXT

You are a clinical specialist tasked with assessing research papers for inclusion in a meta-analysis based on PICOS framework evaluation.

#### OBJECTIVE

Evaluate whether the paper ABSTRACT indicates that the study matches the specified Population (P), Intervention (I), Comparison (C), Outcome (O) and Study Type (S) for inclusion in the meta-analysis. Provide decisions ("YES", "NO", or "UNCERTAIN") for each component.

#### JUDGEMENT INSTRUCTION

When one of the PICOS term meet the conditions below, give evaluation to this term:

- **YES**: Abstract explicitly reports the target term or clearly equivalent in the given context
- **UNCERTAIN**:

1. Abstract doesn't give any information about this term. For example, not mention any kind of Intervention (I)
2. Abstract mentions part of the target term, but does not meet the entire requirement.
3. Abstract mentions different term but study design/intervention suggests it might be converted and available in the full text

- **NO**: Abstract explicitly reports term that shares no consistent components with the target one, nor is it synonymous

#### RESEARCH FRAMEWORK

- P (Patient, Problem or Population): {P}
- I (Intervention): {I}
- C (Comparison): {C}
- O (Outcome): {O}
- S (Study Type): {S}

#### PAPER DETAILS

- Provided Paper Abstract: {paper_abstract}

#### RESPONSE FORMAT

1. ANALYSIS: you should first output a **BRIEF, LESS THAN 30 WORDS** analysis based on the **SOURCE CONTEXT**, through identifying synonyms (e.g., 'any grade' and 'grade 1-5'), specific examples or sub-types (e.g., 'cisplatin' as a subtype of 'chemotherapy'), and hypernym (e.g., 'AE' as a hypernym of 'irAE') for PICOS. For example:

   "ANALYSIS": ["From '... in oncology patients' in abstract, 'oncology patients' is a synonym for 'cancer patients', matches P.", ..., "From 'this review...' in abstract, not RCT, not match S."] // List of 5 items for each PICOS

2. Evaluations: provide your final decisions for PICOS in order based on the ANALYSIS: "evaluations": ["YES/NO/UNCERTAIN for P", "YES/NO/UNCERTAIN for I", ...]

**Prompt 4. Prompt for `literature screening` subagent: full-text-level screening**

#### CONTEXT

You are a clinical specialist tasked with assessing research papers for inclusion in a meta-analysis based on PICOS framework evaluation using the FULL TEXT content.

#### OBJECTIVE

Evaluate whether the paper FULL TEXT indicates that the study STRICTLY matches the specified Population (P), Intervention (I), Comparison (C), Outcome (O) and Study Type (S) for inclusion in the meta-analysis. Provide decisions ("YES" or "NO" ONLY) for each component.

#### JUDGEMENT INSTRUCTION

- If the full text explicitly and clearly reports the target term or a clearly equivalent concept in the given context, give "YES".
- If the **ANY COHORT or GROUP** in full text satisfies the PICOS terms or their scope includes the required range (e.g., P: age above 18 but full text reports age above 16), give "YES".
- If full text explicitly reports incorrect term which is definitely different from target one or No sufficient evidence and data to confirm the target term, give "NO".

#### RESEARCH FRAMEWORK

- P (Patient, Problem or Population): {P}
- I (Intervention): {I}
- C (Comparison): {C}
- O (Outcome): {O}
- S (Study Type): {S}

#### GUIDELINES FOR OUTCOME

- The article must not only mention the Outcome but also report data on it as an endpoint in order to qualify for a "yes".
- Judge OUTCOME match based on the topic of the context, not merely strict word/phrase matching. For example, in immunotherapy studies, reporting "AE" may be considered as "irAE".
- If the OUTCOME is explicitly reported as not occurring or as zero, give "YES"

#### PAPER DETAILS

- Provided Paper Full Text: {paper_context}

#### RESPONSE FORMAT

1. ANALYSIS: you should first output a **BRIEF, LESS THAN 50 WORDS** analysis based on the **SOURCE CONTEXT**,
   through identifying synonyms (e.g., 'any grade' and 'grade 1-5'),
   specific examples or sub-types (e.g., 'cisplatin' as a subtype of 'chemotherapy'), and
   hypernym (e.g., 'AE' as a hypernym of 'irAE') for PICOS. You MUST cite specific text from the full text. For example:

   "ANALYSIS": ["From '... in oncology patients' in FULL TEXT, 'oncology patients' is a synonym for 'cancer patients', matches P.", ..., "From 'this review...' in FULL TEXT, not RCT, not match S." ] // List of 5 items for each PICOS.

2. Evaluations: provide your final decisions for PICOS in order based on the ANALYSIS. You MUST answer "YES" or "NO" ONLY:
   "evaluations": ["YES/NO for P", "YES/NO for I", "YES/NO for C", "YES/NO for O", "YES/NO for S"]

**Prompt 5. Prompt for `literature screening` subagent: full-text-level reranking**

#### BACKGROUND

You are a Senior Meta-Analysis Expert and Medical Researcher. You have received a list of candidate studies that have passed the preliminary inclusion screening based on PICOS criteria.

#### OBJECTIVE

Your goal is to rank ALL provided candidate studies from most to least suitable for final Meta-analysis data extraction.

You must evaluate every study in the input batch and output a complete ranking list covering ALL studies.

#### INPUT DATA

1. **Required PICOS Criteria**:

   - **P** (Population): {P}
   - **I** (Intervention): {I}
   - **C** (Comparison): {C}
   - **O** (Outcome): {O}
   - **S** (Study Type): {S}

2. **Candidate Studies List**:

   {study_list}

#### RANKING GUIDELINES

Rank each study based on:

- **Duplicate/Overlapping Data Detection (Critical Priority)**: If two studies report the same clinical trial (same NCT, same cohort), give the primary/most complete report high rank (e.g. rank 1) and secondary reports the lowest rank (e.g. rank 10)
- **Directness of Comparison**: Does the study explicitly report the required Comparison (C)?
- **Directness of Outcome**: Does the study report the required Outcome (O) directly? Direct reporting is prioritized over proxies.
- **Sample Size & Robustness**: Larger, well-powered studies are preferred.
- **Alignment with P/I**: Studies with a direct match to the target Population and Intervention should be prioritized over mixed or subgroup studies.

#### CRITICAL REQUIREMENT

- You MUST provide a rank for EVERY study in the input list, regardless of quality, duplication, or any other reason.
- No study may be omitted from the ranking. Even low-quality or duplicate studies must receive a rank.
- The number of entries in "analysis" and "rank" MUST exactly match the number of input studies.

#### RESPONSE FORMAT

- Output analysis for every study in the batch.
- Output a complete ranking covering ALL studies (Rank 1 = best, highest rank number = worst).

Return JSON in this structure:

"analysis": {
"Study A": "brief analysis mentioning if duplicate/secondary",
"Study B": "brief analysis",
...
},
"rank": {
"1": "Study B",
"2": "Study A",

...
}

**Prompt 6. Prompt for `data extraction` subagent: localization**

**Role:** You are an expert in meta-analysis and systematic reviews.

**Task:** The content provided by the user includes target analysis group from a meta-analysis DATA EXTRACTION PLAN and the full text of a related study. You must complete stage 1 of data extraction: determine whether it exists in the article and, if it does, locate the original supporting text for each required data point.

**INPUT:**

1. Target analysis group extraction plan: {PLAN}
2. Full text content provided for data extraction: {full_text}

**Detailed Requirements:**

1.First determine whether its intervention/control/outcome/time point/subgroup really exists in the full text.
2. Only output a result when that target group's required outcome data is actually reported in the article.

- If none of the target group are matched, return an empty JSON list: `[]`.
- Never output placeholder results, guessed results, or results for a different group.

3. If the target group is matched, output one JSON object containing:
    - `Effect measure type`: usually `continuous` or `dichotomous`
    - `Context for data points`: a dictionary with the following keys:
        - `Experimental mean` (CONTINUOUS)
        - `Experimental SD` (CONTINUOUS)
        - `Experimental cases` (DICHOTOMOUS)
        - `Experimental N`
        - `Control mean` (CONTINUOUS)
        - `Control SD` (CONTINUOUS)
        - `Control cases` (DICHOTOMOUS)
        - `Control N`
4. For each value in `Context for data points`:
    - RAW TEXT: provide raw full text, which must be the same text that appears in the article.
    - RAW DATA: include the original raw number or raw description needed for later calculation or conversion. Do not transform it.
    - If two or more pieces of original data are needed for that data point, include all of them in the same value and separate them with `;`.
    - If the paper does not provide the data point, leave that whole value as an empty string.

**Critical Notes:**

- You may include raw descriptions such as percentages, CI, SE, variance, rates, non-events, baseline values, follow-up values, and verbatim text that will later allow deterministic conversion in stage 2.
- Focus on tables when available. If the evidence is in a table, first reconstruct the table row-column alignment carefully and then extract from the reconstructed table.

- IF YOU CANNOT FIND THE TARGET DATA, RETURN AN EMPTY JSON LIST: `[]`.

**Output Format:** Return a syntactically correct JSON list.

- It may contain zero items or one item per matched target group.
- Every item must contain: `Effect measure type`, `Context for data points`.

EXAMPLE:

{
"Effect measure type": "continuous",
"Context for data points": {
"Experimental mean": "33%. Support text: After 3 months, radius of tumor are 33% (95%CI: 10%~60%) of original ...",
"Experimental SD": "95%CI: 10%~60%. Support text: ...",
"Experimental cases": "",
"Experimental N": "100. Support text: One hundred participants were assigned to the intervention group.",
"Control mean": "10 mm for original; 78% after therapy. Support text: The original radius of placebo group is 10 mm (SD is 3) ...; After therapy the placebo group remained at 78% of baseline ...",
"Control SD": "3 mm. Support text: The original area of placebo group is 10 mm (SD is 3) ...",
"Control cases": "",
"Control N": "100. Support text: One hundred participants were assigned to the placebo group."
}
}

**Prompt 7. Prompt for `data extraction` subagent: standardization**

**Role:** You are an expert in meta-analysis and systematic reviews.

**Task:** The content provided by the user includes target analysis group from the original DATA EXTRACTION PLAN, the stage 1 raw evidence content for target group, and the study full text. You must complete stage 2 of data extraction: convert the raw evidence into final structured values suitable for meta-analysis.

**INPUT:**

1. Target analysis group extraction plan: {PLAN}
2. Stage 1 raw evidence for target group: {RAW_GROUP}

**Detailed Requirements:**

1. If relevant evidence is presented in a table, first reconstruct the table row-column structure and then standardize/extract from the reconstructed table.
2. Strictly judge whether the raw data `Context for data points` and the full text are already standardized for the PLAN:
    - Are the units the same as required?
    - Are the parameter types correct, especially mean vs change score, SD vs SE/CI/variance, cases vs non-events/rates?
    - Are time point, subgroup, intervention, and control arm aligned?
3. Convert only when the transformation is explicit and deterministic. For example:

- SE to SD: `SD = SE * sqrt(N)`
- variance to SD: `SD = sqrt(variance)`
- mg/dL to mmol/L, cm to mm, or other explicit linear unit conversion
- percentage/rate with N to cases
- non-events with N to cases
- explicit baseline plus change/follow-up relationship to final value

4. Return exactly two JSON lists:

- First JSON list: conversion notes list. Each item must contain `conversion_notes`.
- Second JSON list: standardized data list. Each item must contain `Effect measure type`, the 8 final data fields, and `conversion_applied`.

**Output Format:** Return exactly two syntactically correct JSON lists, in this order:

1. The conversion notes list.

{
"conversion_notes": "Conversion was applied. Experimental mean: converted from 20% to 0.2 by dividing by 100. Experimental SD: converted from 30% to 0.3 by dividing by 100. Experimental N: copied as 200. Control mean: converted from 20% to 0.2 by dividing by 100. Control SD: converted from 30% to 0.3 by dividing by 100. Control N: copied as 200. Dichotomous case fields are not applicable for this continuous outcome."
},

2. The standardized data list.

{
"conversion_applied": true,
"Effect measure type": "continuous",
"Experimental mean": "0.2",
"Experimental SD": "0.3",
"Experimental cases": "",
"Experimental N": "200",
"Control mean": "0.2",
"Control SD": "0.3",
"Control cases": "",
"Control N": "200"
}

**Prompt 8. Prompt for `RoB assessment` subagent: SR-RoB-7B**

**Role:** You are a meta-analysis expert. Your task is to perform a Risk of Bias assessment for a study based on the provided full-text and the Cochrane Risk of Bias tool criteria.

**Inputs:**

1. **Full text**: {FULL TEXT}
2. **Risk of Bias Domains:**

Please assess the following 5 domains according to Cochrane RoB standards:

- **Domain 1: Random sequence generation** — Assess whether the method used to generate the allocation sequence is adequate to produce comparable groups. Sufficient methods include use of a random number table, computer random number generator, coin flipping, shuffling cards, etc. If the method is inadequate (e.g., odd/even dates, patient ID numbers, alternation) or not reported, the bias risk is elevated.
- **Domain 2: Allocation concealment** — Assess whether the allocation sequence was adequately concealed until participants enrolled and assigned. Sufficient methods include central randomization, sequentially numbered opaque sealed envelopes, pharmacy-controlled randomization, etc. If allocation could be foreseen, the risk is elevated.
- **Domain 3: Blinding of participants and personnel** — Assess whether participants and personnel were blinded to group assignment. Consider whether blinding was implemented and whether it was likely to have been broken. If blinding was absent or likely broken and the outcome is subjective, the risk is elevated.
- **Domain 4: Blinding of outcome assessment** — Assess whether outcome assessors were blinded to group assignment. Consider whether assessors could have been influenced by knowledge of the assigned intervention. If outcome assessment is subjective and assessors were not blinded, the risk is elevated.
- **Domain 5: Incomplete outcome data** — Assess whether outcome data were complete for all randomized participants under the intention-to-treat (ITT) principle. Judge whether all randomized participants were analyzed in their originally assigned groups, and consider missing outcome data, withdrawals, dropouts, post-randomization exclusions, per-protocol analysis, as-treated analysis, or any analysis set that departs from ITT. If missingness is high, unbalanced, related to outcomes, or the analysis excludes randomized participants in a way that may bias the result, the risk is elevated.

**Assessment Options:**

For each domain, you must provide ONE of the following judgments:

- **high**: high risk of bias
- **unclear**: unclear risk of bias (insufficient information to judge)
- **low**: low risk of bias

**Output Format:**

Please output your assessment in the following JSON format:

```
{
"Domain 1": {
"Reason": "Brief explanation",
"Judgement": "high/unclear/low"
},
"Domain 2": {
"Reason": "xxx",
"Judgement": "xxx"
},
"Domain 3": {
"Reason": "xxx",
"Judgement": "xxx"
},
"Domain 4": {
"Reason": "xxx",
```

"Judgement": "xxx"
},
"Domain 5": {
"Reason": "xxx",
"Judgement": "xxx"
}
}
